# Multi-modal characterisation of cardiac function and electrophysiology in type 2 diabetes: a UK Biobank cross-sectional study

**DOI:** 10.1101/2024.06.26.24309474

**Authors:** Ambre Bertrand, Andrew Lewis, Julia Camps, Vicente Grau, Blanca Rodriguez

## Abstract

**Background and Aim:** Type 2 diabetes mellitus (T2DM) is a major risk factor for heart failure, ischemic heart disease, and cardiac arrhythmias. Our goal is to examine the association of T2DM with ECG and cardiac imaging biomarkers, providing a window into the adverse effects of T2DM on cardiac health.

**Methods:** Using data from the UK Biobank, we investigated ECG and cardiac magnetic resonance imaging biomarkers in a cohort of 1781 participants with T2DM and no diagnosed cardiovascular disease at time of assessment. We performed a pair-matched cross-sectional study to examine the association between type 2 diabetes and multi-modal cardiac biomarkers. We built multivariate multiple linear regression models sequentially adjusted for socio-demographic, lifestyle, and clinical covariates.

**Results:** T2DM was associated with a higher resting heart rate (66 vs 61 beats per minute, p<0.001), longer QTc interval (424 vs 420 ms, p<0.001), reduced T-wave amplitude (0.33 vs 0.37 mV, p<0.001), lower stroke volume (72 vs 78 ml, p<0.001) and thicker left ventricular wall (6.1 vs 5.9 mm, p<0.001). These trends were consistent in subgroups of different sex, age and body mass index. Fewer significant differences were noted in non-white participants. QRS duration and Sokolow-Lyon index were associated with the development of cardiovascular disease in groups with and without T2DM, respectively. A higher left ventricular mass and wall thickness were associated with cardiovascular outcomes in both groups.

**Conclusion:** T2DM was associated with adverse changes in ECG and cardiac imaging biomarkers, possibly reflecting subclinical cardiac repolarisation abnormalities, autonomic dysfunction, hypertrophy and impaired mechanical function.

## Introduction

Type 2 diabetes mellitus (T2DM) is a complex metabolic disease that affects over 536 million adults worldwide, according to the International Diabetes Federation (IDF) [1]. T2DM causes a two-to four-fold increase in risk of cardiovascular disease (CVD), the leading cause of deaths globally, which claims the lives of 17.9 million people annually according to the World Health Organisation [2,3]. Thus, there is a pressing need to better understand and identify the adverse effects of T2DM on the cardiovascular system, in order to mitigate disease progression and ultimately reduce deaths due to T2DM-driven CVD.

T2DM is characterised by chronically elevated blood glucose levels, known as hyperglycaemia, due to insulin resistance. Hyperglycaemia triggers a series of adverse molecular changes that lead to myocardial fibrosis, stiffness, and contractile dysfunction. These changes worsen progressively over time, eventually resulting in diastolic dysfunction and heart failure if left uncontrolled. Additionally, diabetes is associated with electrophysiological changes caused by the modulation of cardiac ionic currents, alterations in gap junctions, and abnormal conduction due to fibrosis [4–6]. Cardiac autonomic neuropathy (CAN), a common yet underdiagnosed complication of diabetes, is also responsible for altering electrophysiological function and heart rate due to damaged cardiac innervation [7]. Cardiac magnetic resonance (CMR) imaging has identified structural and functional changes in individuals with diabetes of unspecified type [8], while electrophysiological abnormalities in diabetes may manifest in the electrocardiogram (ECG), notably in the QT interval and T wave [9–12].

However, to our knowledge, no previous work has quantified ECG and CMR-derived biomarkers concurrently in a large prospective cohort of individuals with diabetes of type 2 specifically, before clinical diagnosis of CVD. The goal of this study is to improve our understanding and the identification of subclinical T2DM-driven cardiac remodelling at a population level, ultimately supporting CVD risk stratification in patients with T2DM. We hypothesise that, compared to matched controls, ECG and CMR-derived biomarker differences in individuals with T2DM and no prior cardiovascular events may uncover signs of diabetic cardiomyopathy and subclinical electrophysiological abnormalities. Using the UK Biobank, we investigate the effect of T2DM on multi-modal biomarkers reflecting cardiac structure, function and electrophysiology, and assess these biomarkers’ predictive value in relation to CVD development. Sex, age, body mass index, and ethnicity-related differences are investigated using subgroup analyses, as well as differences across the glycaemic spectrum.

## Methods

### Study design and population

The UK Biobank study is a multi-centre, prospective cohort study of over half a million adults recruited between the age of 40 and 69, and living in England, Scotland and Wales. It contains socio-demographic, lifestyle, and clinical information recorded at multiple timepoints since recruitment in 2006, and is linked to general practice primary care records and hospital episode statistics (HES).

Following a baseline assessment visit attended by all participants, about 50,000 participants selected at random were recalled for a multi-modal imaging assessment. This visit included a CMR scan performed on a 1.5 Tesla scanner (MAGNETOM Aera, Syngo Platform VD13A, Siemens Healthcare). Full details of the imaging protocol are available in [13]. A 12-lead resting ECG (CardioSoft ECG system, GE) was also recorded on the same day. Our primary analysis is a matched cross-sectional study examining the association between T2DM status and multi-modal cardiac biomarkers. As secondary analyses, we also carry out a case-control study quantifying the relationship between selected biomarkers in participants who did and did not develop CVD during follow-up, in two matched cohorts with and without T2DM.

Our study is reported in line with the STROBE (Strengthening the Reporting of Observational Studies in Epidemiology) statement.

### Ethical considerations

General ethical approval was granted for UK Biobank studies by the United Kingdom’s National Health Service Research Ethics Service (11/NW/0382). Participants provided written informed consent for their data to be stored and used for research purposes. This study was conducted under UK Biobank Application Number 40161.

### Cohort definition

### Disease ascertainment

The selection process and cohort sizes are summarised in Figure 1. We defined our cohorts based on the clinical ascertainment of two disease phenotypes, namely T2DM and CVD. Relevant ICD-9 and ICD-10 codes and their corresponding UK Biobank fields are listed in Supplementary Table 1. T2DM was determined by one or more of the following criteria: date of a T2DM ICD code first reported, HES record corresponding to a T2DM ICD code, HbA1c ≥ 48 mmol/mol, response to a patient touchscreen questionnaire [14]. CVD was determined by one or more of the following criteria: date of a CVD ICD code first reported, HES record corresponding to a CVD ICD code, date of myocardial infarction (MI), date of ST-elevated MI, and date of non-ST-elevated MI. The last three data fields are UK Biobank-specific algorithmically defined diagnoses of MI summarising information contained in other fields; cases identified via those fields may overlap with other routes of ascertainment but were included for completeness.

**Figure 1.**
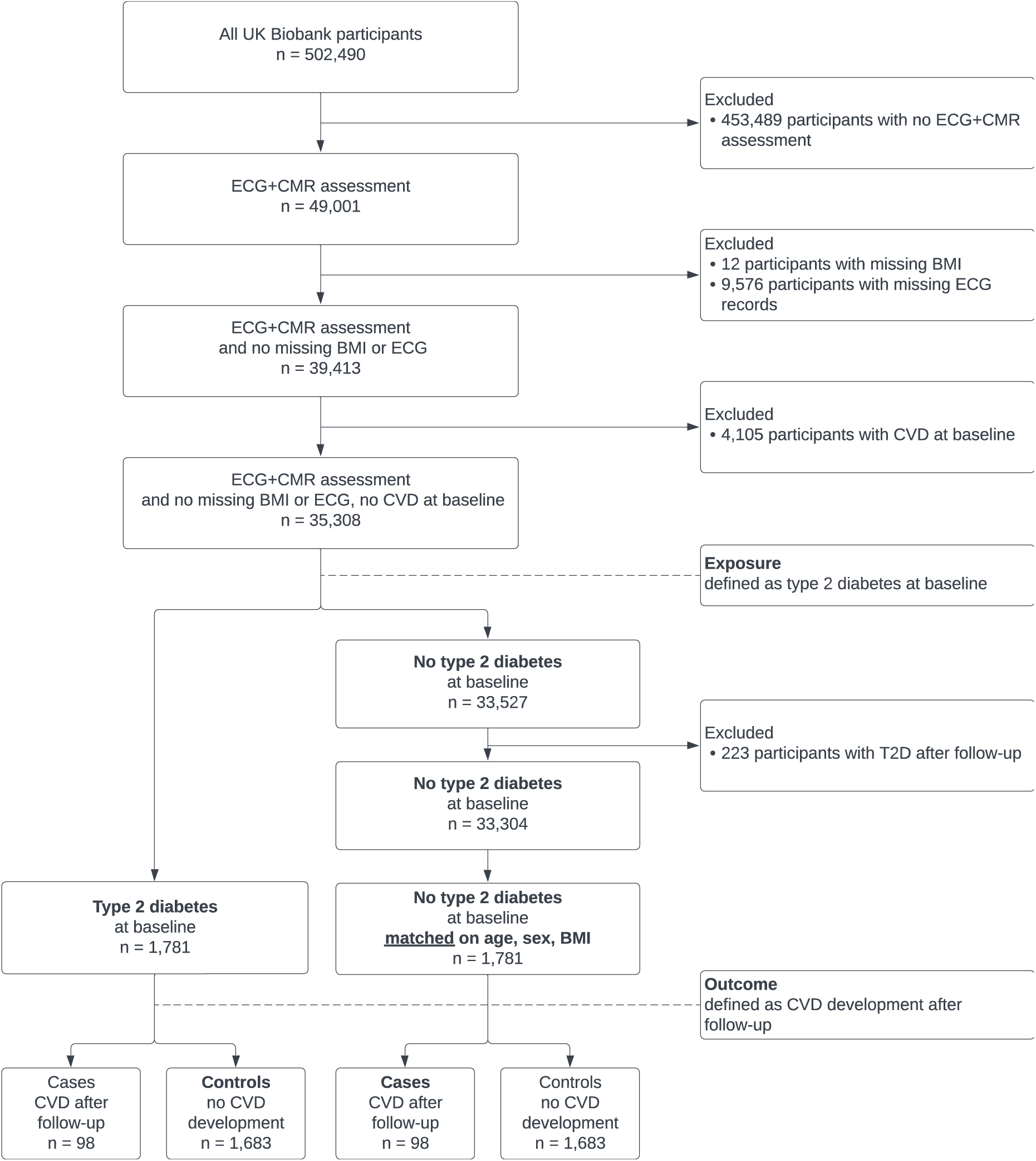
Cohort selection flowchart. CMR: cardiac magnetic resonance imaging, BMI: body mass index, CVD: cardiovascular disease, T2D: type 2 diabetes. Baseline refers to the time of recording of the ECG and CMR images. Follow-up refers to the period between baseline and censoring date (21/09/2021).

### Exclusion criteria, exposure and outcomes

We excluded all participants with pre-existing CVD, and censored all participants with CVD diagnosed after 21/09/2021, the date of the latest CVD diagnosis identified at the time of analysis. The median follow-up period between date of assessment and date of first CVD diagnosis is 1.5 years (min 8 days, max 5.9 years). The breakdown of follow-up CVDs in each cohort is presented in Supplementary Figure 1.

The exposure cohort consists of eligible participants with known T2DM at time of assessment. Pair-matching was carried out to select controls with no known T2DM based on age, sex, body mass index (BMI), as well as binary occurrence of CVD during follow-up (i.e. with vs without a CVD diagnosis). We use BMI as a suitable matching variable as it is a commonly used marker of adiposity that modulates the ECG [15]. Euclidian distances were computed between the [age, BMI] vectors of each exposure participant and possible control participants. Each exposure participant was matched by a control participant of the same sex and follow-up CVD status with the lowest [age, BMI] Euclidian distance [16].

We defined outcomes as ECG and CMR biomarker values for all analyses apart from a secondary analysis considering CVD development, where we defined an outcome as a case of CVD during follow-up.

### Baseline covariates

### Demographic, lifestyle, and clinical characteristics

We included the following demographic, lifestyle, and clinical characteristics: age at assessment, sex, ethnicity, weight, height, smoking status, systolic blood pressure (SBP), diastolic blood pressure (DBP), medication. BMI was calculated using the ratio of weight in kilograms over squared height in meters. Information on medication was self-reported and divided into broad categories and/or using UK Biobank-specific individual medication codes recorded by nurses. We included cholesterol-lowering medication, blood pressure medication, insulin, metformin and rosiglitazone 1mg/metformin 500mg tablet. The following blood biochemistry measurements were included: high-density lipoprotein (HDL) cholesterol, total cholesterol, HbA1c, triglycerides, creatinine, C-reactive protein. These measurements were taken prior to the imaging assessment so we used the most recent one available. Estimated glomerular filtration rate (eGFR) was calculated using serum creatinine according to the CKD-EPI creatinine equation adjusted for age, sex and ethnicity [17]. Creatinine was converted to mg/dL by multiplying the given value in µmol/L by a factor of 0.0113 [18].

### ECG and CMR-derived biomarkers

Resting ECG recordings were available in .xml file format. Each file included the raw ECG signal as well as automatically extracted key ECG markers. Wave amplitudes were available for all 12 leads, while other markers were available for lead I only. We used the following markers: ventricular rate, QRS duration, QTc interval, T wave offset, R-wave amplitude, S-wave amplitude, J point amplitude (equivalent to ST segment elevation or depression), and T wave amplitude. Pairwise Pearson correlation coefficients were calculated to assess inter-covariate correlations among T wave and J point amplitudes in different ECG leads (Figure 2). Two representative leads were retained, one limb lead (aVL) and one precordial lead (V3). We computed the Sokolow-Lyon index for left ventricular hypertrophy as the sum of the S amplitude in V1 and the maximum value of R amplitude in V5 or V6 [19].

**Figure 2.**
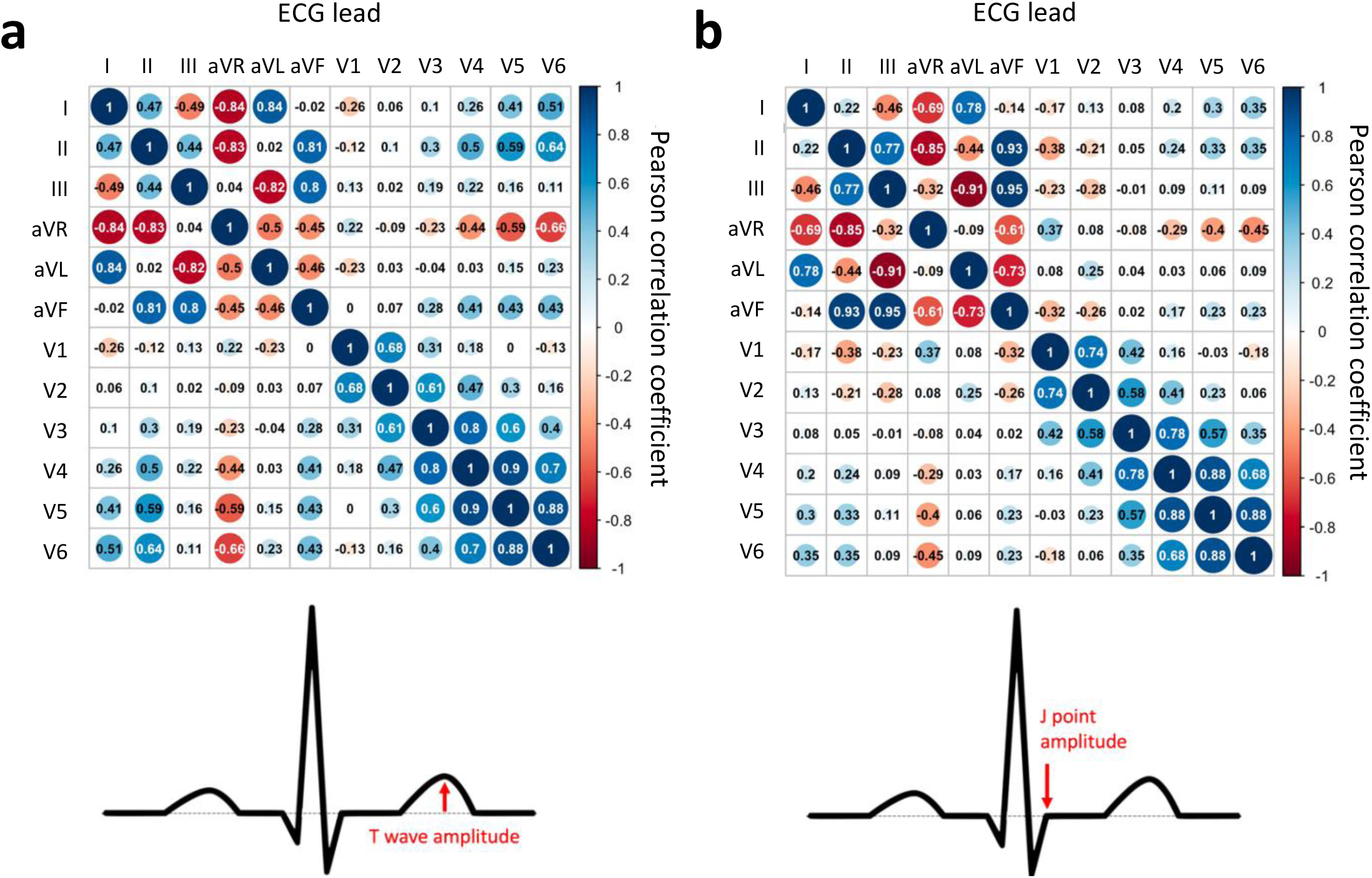
Matrix of Pearson correlation coefficients for 12-lead ECG amplitude measurements of T wave amplitude (a) and J point amplitude (b). Coefficients were computed on 2702 (a) and 3562 cases (b), respectively. These cases contained no missing data among the features of interest.

Some key CMR-derived biomarkers were directly available in the UK Biobank database. Manual analyses, algorithms and validation of these biomarkers are described in detail in [20,21]. We used the following in our study: left ventricular ejection fraction (LVEF), LV end-diastolic volume, LV end-systolic volume, LV stroke volume, cardiac output, LV myocardial mass, LV global average wall thickness.

### Statistical analysis

All statistical analyses were performed using RStudio with R version 4.1.1.

#### Comparison of ECG and CMR biomarkers in individuals with and without type 2 diabetes

Baseline covariates were compared between the exposure and control cohorts. Normality of continuous variable distributions were assessed using the Shapiro-Wilk test. The independent samples t-test was used for normal distributions and the Mann-Whitney U test was used for non-normal distributions. Missing data were not imputed. All statistical tests were two-sided. Statistical significance was defined as p<0.05. Median, interquartile range (IQR) and missing measurements in the form of count and percentage of the full sample, are reported for each variable.

### Association of type 2 diabetes with ECG and CMR biomarkers

We used multivariate multiple linear regression to examine the association between T2DM and ECG and CMR biomarkers. A crude, unadjusted model (Model 0) was used to quantify the association of T2DM only with each biomarker. Three additional models were constructed incrementally to account for socio-demographic (Model 1), lifestyle (Model 2), and clinical covariates detailed in the previous section (Model 3; fully adjusted). Pairwise Pearson correlation coefficients were calculated to assess inter-covariate correlation for all potential model covariates. Coefficients >0.3 or <-0.3 determined the exclusion of the following covariates: eGFR, HbA1c, systolic blood pressure, HDL cholesterol, cholesterol-lowering medication (Supplementary Figure 2). Models were fitted using the Ordinary Least Squares method, therefore scaling of variables was not required. Regression coefficients, their 95% confidence intervals, and p-values are reported for each outcome variable and model in the tables.

### Type 2 diabetes and occurrence of cardiovascular disease

We compared biomarkers between cases who did and did not develop CVD, in both cohorts with and without T2DM. We assessed the association between biomarkers with significant differences, and CVD development. We built two sets of logistic regression models, one for each cohort, and examined how these associations differ in each cohort. We followed the same covariate processing and sequential adjustment approach as described previously.

### Subgroup analyses

We also examined the role of sex, age, BMI and ethnicity by comparing biomarkers in stratified subgroups. We investigated the effect of severity of disease on selected biomarkers by considering HbA1c as a proxy for hyperglycaemia. We built regression models as described previously, this time using HbA1c as a continuous exposure variable, and biomarkers that were associated with T2DM as outcomes. This analysis was performed within the T2DM cohort only, to avoid a dichotomous HbA1c distribution due to lower HbA1c levels in the control cohort.

## Results

### Cohort description

Both the exposure and the control cohorts (T2DM vs no T2DM) consisted of 1,781 subjects matched by age, sex and BMI. Cohorts were mostly male (n=1133, 63.6%), had a median age of 67 years and a median BMI of 27.8 kg/m^2^, which is considered overweight by the NHS (NHS Digital, 2022) (Table 1). Individuals with T2DM were more likely to be non-white (7.9% vs 2.7%) and to be current or previous smokers (43.5% vs 40.6%). They tended to have lower diastolic blood pressure, total cholesterol, and HDL cholesterol, while HbA1c, triglycerides, C-reactive protein, and eGFR were generally more elevated. Subjects with T2DM were also more likely to be on cholesterol-lowering drugs, anti-hypertensives, insulin and metformin. One patient in the control cohort reported taking insulin and metformin. This may be due to a lack of diabetes diagnosis at the time of visit, or an increase in HbA1c levels after the last measurement, leading to a missed positive classification of disease. Although rare, this case could also be due to the medication being used to treat another condition unrelated to diabetes, such as polycystic ovary syndrome [22].

**Table 1.**
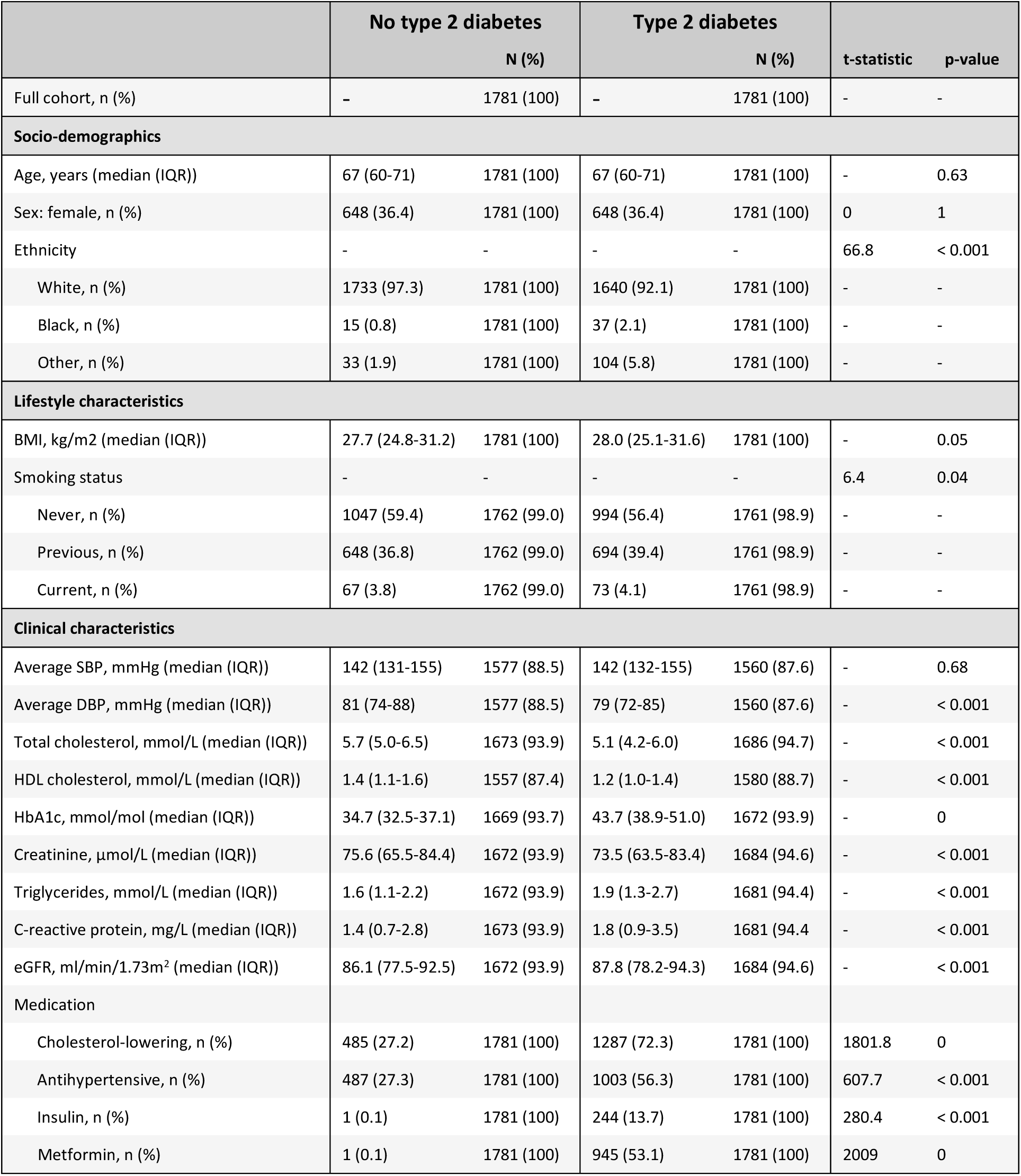
Socio-demographic, lifestyle, and clinical characteristics recorded for the cohorts with and without type 2 diabetes. IQR stands for inter-quartile range; BMI, body mass index; SBP, systolic blood pressure; DBP, diastolic blood pressure; HDL, high-density lipoprotein; HbA1c, glycated haemoglobin; eGFR, estimated glomerular filtration rate. The t-statistic is reported for categorical variables assessed using the Chi-Squared test. All continuous variables are distributed non-normally and compared using the Mann-Whitney U-test.

### Type 2 diabetes and ECG biomarkers

Compared to matched controls, ventricular rate in T2DM was higher (+5 bpm, median: 66 IQR: [59–74] vs 61 [55–68] bpm) and T wave offset was earlier (-12 ms, 842 [820–864] vs 854 [834–874] ms) (Table 2). T2DM was strongly associated with both biomarkers in all models (Table 3). This consistency is expected, given the known inverse correlation between T wave offset and ventricular rate. QRS duration was shorter (-2 ms, 86 [79–93] vs 88 [82–96] ms) and QTc interval was longer (+4 ms, 424 [408–440] vs 420 [405–436] ms) in the T2DM cohort (Table 2). There was a statistically significant association of T2DM with QTc interval in all models but the fully adjusted one (Table 3). However, the association of T2DM with QRS duration was significant only in the two last models adjusted for lifestyle and clinical factors (Model 2 and 3, Table 3). This suggests that the importance of this variable may increase relative to other covariates included in the later models. There were statistically significant differences in almost all wave amplitude biomarkers between the cohorts (Table 2). T2DM was associated with a flatter T wave amplitude in V3 in all models; a flatter T wave amplitude and less elevated J-point in aVL in all models but the fully adjusted one, and a less elevated J-point in aVL (Table 3). T2DM was associated with a lower Sokolow-Lyon index in all models but the fully adjusted one (Tables 2 and 3).

**Table 2.** ECG and CMR-derived biomarkers recorded for the cohorts with and without type 2 diabetes. IQR stands for inter-quartile range; ECG, electrocardiogram; CMR, cardiac magnetic resonance; LV, left ventricular; EF, ejection fraction; ED, end diastolic; ES, end systolic. All continuous variables are distributed non-normally and compared using the Mann-Whitney U-test.

|  | No type 2 diabetes |  | Type 2 diabetes |  | p-value |
| --- | --- | --- | --- | --- | --- |
|  | Median (IQR) | N (%) | Median (IQR) | N (%) |  |
| Full cohort | - | 1781 (100) | - | 1781 (100) | - |
| **ECG** |  |  |  |  |  |
| Ventricular rate, bpm | 61 (55-68) | 1781 (100) | 66 (59-74) | 1781 (100) | < 0.001 |
| QRS duration, ms | 88 (82-96) | 1781 (100) | 86 (79-93) | 1781 (100) | 0.005 |
| QTc interval, ms | 420 (405-436) | 1781 (100) | 424 (408-440) | 1781 (100) | < 0.001 |
| T-wave offset, ms | 854 (834-874) | 1781 (100) | 842 (820-864) | 1781 (100) | < 0.001 |
| T-wave amplitude (V3), mV | 0.37 (0.23-0.54) | 1766 (99.2) | 0.33 (0.21-0.49) | 1765 (99.1) | < 0.001 |
| T-wave amplitude (aVL), mV | 0.11 (0.05-0.17) | 1718 (96.5) | 0.10 (0.04-0.15) | 1669 (93.7) | < 0.001 |
| J-point amplitude (V3), mV | -0.02 ((-0.04)-0.02) | 1781 (100) | -0.02 ((-0.04)-0.02) | 1781 (100) | 0.89 |
| J-point amplitude (aVL), mV | 0.01 ((-0.01)-0.03) | 1781 (100) | 0.00 ((-0.02)-0.03) | 1781 (100) | 0.016 |
| Sokolow-Lyon index, mm | 20.2 (16.2-24.8) | 1650 (92.6) | 19.1 (15.2-23.5) | 1612 (90.5) | < 0.001 |
| **CMR** |  |  |  |  |  |
| LVEF, % | 56 (52-59) | 1533 (86.1) | 55 (51-59) | 1501 (84.3) | 0.066 |
| LV ED volume, ml | 140 (119-163) | 1533 (86.1) | 130 (109-155) | 1501 (84.3) | < 0.001 |
| LV ES volume, ml | 61 (51-74) | 1533 (86.1) | 58 (47-71) | 1501 (84.3) | < 0.001 |
| LV stroke volume, ml | 78 (65-90) | 1533 (86.1) | 72 (60-85) | 1501 (84.3) | < 0.001 |
| Cardiac output, ml | 4.7 (4.1-5.5) | 1533 (86.1) | 4.7 (4.0-5.5) | 1501 (84.3) | 0.53 |
| LV mass, g | 92 (76-109) | 1518 (85.2) | 91 (77-108) | 1518 (85.2) | 0.87 |
| LV global average wall thickness, mm | 5.9 (5.4-6.5) | 1516 (85.1) | 6.1 (5.6-6.6) | 1516 (85.1) | < 0.001 |

**Table 3.** Multivariate multiple linear regression models used to quantify the association of type 2 diabetes with selected ECG and CMR-derived biomarkers. Models are adjusted sequentially for different types of confounding factors. Socio-demographic factors include age, sex, ethnicity; lifestyle factors include body mass index (BMI), smoking; clinical factors include diastolic blood pressure, total cholesterol, triglycerides, C-reactive protein, anti-hypertensive medication and insulin.

| Outcome | Model 0<br>Unadjusted<br>N = 2577 |  | Model 1<br>Adjusted for socio-demographic factors<br>N = 2577 |  | Model 2<br>Additionally adjusted for lifestyle factors<br>N = 2544 |  | Model 3<br>Additionally adjusted for clinical factors<br>N = 2201 |  |
| --- | --- | --- | --- | --- | --- | --- | --- | --- |
|  | Coefficient<br>(95% CI) | p-value | Coefficient<br>(95% CI) | p-value | Coefficient<br>(95% CI) | p-value | Coefficient<br>(95% CI) | p-value |
| **ECG** |  |  |  |  |  |  |  |  |
| Ventricular rate, bpm | 4.14<br>(3.30-4.98) | <0.001 | 4.15<br>(3.31-4.98) | <0.001 | 3.84<br>(3.01-4.67) | <0.001 | 3.11<br>(2.11-4.1) | <0.001 |
| QRS duration, ms | -0.76<br>(-1.81-0.28) | 0.151 | -0.89<br>(-1.89-0.10) | 0.079 | -1.14<br>(-2.14-(-0.14)) | 0.026 | -1.81<br>(-3.01-(-0.61)) | 0.003 |
| QTc interval, ms | 2.57<br>(0.78-4.37) | 0.005 | 2.48<br>(0.76-4.20) | 0.005 | 1.83<br>(0.13-3.53) | 0.035 | 0.86<br>(-1.22-2.95) | 0.416 |
| T-wave offset, ms | -10.5<br>(-12.9-(-8.0)) | <0.001 | -10.6<br>(-13.0-(-8.2)) | <0.001 | -10.1<br>(-12.5-(-7.6)) | <0.001 | -8.3<br>(-11.2-(-5.3)) | <0.001 |
| T-wave amplitude (V3), mV | -0.04<br>(-0.06-(-0.02)) | <0.001 | -0.038<br>(-0.06-(-0.02)) | <0.001 | -0.031<br>(-0.048-(-0.014)) | <0.001 | -0.025<br>(-0.047-(-0.004)) | 0.021 |
| T-wave amplitude (aVL), mV | -0.01<br>(-0.02-(-0.01)) | <0.001 | -0.01<br>(-0.02-(-0.01)) | <0.001 | -0.01<br>(-0.02-(-0.01)) | <0.001 | -0.005<br>(-0.015-0.004) | 0.285 |
| J-point amplitude (V3), mV | 0.002<br>(-0.003-0.006) | 0.437 | 0.002<br>(-0.003-0.006) | 0.47 | 0.003<br>(-0.001-0.007) | 0.198 | 0.003<br>(-0.002-0.008) | 0.218 |
| J-point amplitude (aVL), mV | -0.004<br>(-0.007-(-0.001)) | 0.007 | -0.004<br>(-0.006-(-0.001)) | 0.009 | -0.003<br>(-0.006-(-0.001)) | 0.019 | -0.001<br>(-0.004-0.002) | 0.573 |
| Sokolow-Lyon, mm | -0.83<br>(-1.34-(-0.32)) | 0.001 | -0.80<br>(-1.29-(-0.30)) | 0.002 | -0.61<br>(-1.09-(-0.12)) | 0.015 | -0.39<br>(-0.99-0.21) | 0.199 |
| **CMR** |  |  |  |  |  |  |  |  |
| LVEF, % | -0.415<br>(-0.952-0.123) | 0.131 | -0.371<br>(-0.902-0.161) | 0.171 | -0.289<br>(-0.825-0.247) | 0.29 | -0.195<br>(-0.839-0.449) | 0.553 |
| LV ED volume, mL | -8.63<br>(-15.2-(-2.09)) | 0.010 | -8.44<br>(-14.8-(-2.03)) | 0.010 | -9.11<br>(-15.6-(-2.61)) | 0.006 | -6.86<br>(-15.3-1.55) | 0.11 |
| LV ES volume, mL | -3.47<br>(-9.09-2.15) | 0.226 | -3.48<br>(-9.06-2.1) | 0.221 | -3.95<br>(-9.62-1.71) | 0.171 | -2.79<br>(-10.2-4.59) | 0.459 |
| LV stroke volume, mL | -5.18<br>(-6.87-(-3.5)) | <0.001 | -4.98<br>(-6.53-(-3.42)) | <0.001 | -5.17<br>(-6.74-(-3.6)) | <0.001 | -4.11<br>(-6.03-(-2.19)) | <0.001 |
| LV cardiac output, L/min | -0.0517<br>(-0.158-0.055) | 0.341 | -0.038<br>(-0.137-0.061) | 0.453 | -0.0622<br>(-0.161-0.037) | 0.219 | -0.0429<br>(-0.165-0.080) | 0.493 |
| LV mass, g | 0.359<br>(-1.38-2.1) | 0.687 | 0.466<br>(-0.846-1.78) | 0.487 | -0.388<br>(-1.59-0.811) | 0.525 | -0.165<br>(-1.63-1.3) | 0.825 |
| LV global average wall thickness, mm | 0.173<br>(0.113-0.232) | <0.001 | 0.174<br>(0.125-0.223) | <0.001 | 0.139<br>(0.094-0.183) | <0.001 | 0.133<br>(0.081-0.186) | <0.001 |

### Type 2 diabetes and CMR-derived biomarkers

Left ventricular wall thickness was higher in the T2DM cohort (+2 mm, 6.1 (5.6-6.6) vs 5.9 (5.4-6.5) mm) (Table 2), and its association with T2DM was statistically significant in all models (Table 3). Left ventricular stroke volume was lower in the T2DM cohort (-6 ml, 72 (60-85) vs 78 (65-90) ml), as were end-diastolic and end-systolic volumes (Table 2). However, the association of T2DM was significant in all models only with stroke volume (Table 3). No statistically significant differences or associations were observed for LVEF nor cardiac output.

### Type 2 diabetes and cardiovascular disease outcomes

T2DM individuals who went on to develop CVD (n=98) compared to those who did not (n=1683) were more likely to be male (70% vs 64%) and more likely to take metformin (57% vs 53%). In this cohort of subjects with diabetes, there were no changes observed in blood pressure, cholesterol, HbA1c, or eGFR between individuals who do and do not develop CVD. In the T2DM cohort, cases who developed CVD during follow-up (n=98) had a longer QRS duration (median: 88 ms, IQR: [82–98] vs 86 [80–94] ms, p=0.03) (Supplementary Table 2). There was a statistically significant association between QRS duration and CVD development in all models but the fully adjusted one (Supplementary Table 3). In subjects without T2DM, cases of CVD (n=98) had a higher Sokolow-Lyon index (23.4 [17.7-27.1] vs 20.1 [16.2-24.7] mm, p=0.005). We found a statistically significant association between the Sokolow-Lyon index and CVD development in all models (Supplementary Table 3). In both cohorts, cases with CVD had a higher left ventricular mass (T2DM: 100 [86–107] vs 91 [76–108] g, p=0.006; no T2DM: 101 [83–118] vs 92 [76–108] g, p<0.001) and a thicker left ventricular wall (T2DM: 6.3 [5.9-6.8] vs 6.1 [5.6-6.6] mm, p=0.006; no T2DM: 6.3 [5.6-6.9] vs 5.9 [5.4-6.4] mm, p<0.001) (Supplementary Table 2). Both variables showed a statistically significant association with CVD in all models (Supplementary Table 3).

### Subgroup analyses

### Sex, age, and body mass index

Similar trends were observed between biomarkers of subjects with and without T2DM, in females (n=648) and males (n=1133). Compared to those without T2DM, both females and males in the T2DM group exhibited increased ventricular rates, a prolonged QTc interval, an earlier T wave offset, flatter T wave amplitudes in leads V3 and aVL, a lower Sokolow-Lyon index, lower end-diastolic, end-systolic, and stroke volumes, and a thicker left ventricular wall (Supplementary Table 5). As expected, baseline biomarker values are different in males and females. Sex-specific results stratified by age and BMI are available in Supplementary Figures 3 and 4. No notable differences were observed across age groups. In different BMI groups, there was a gradual decrease in Sokolow-Lyon index with increasing BMI in both cohorts, likely explained by increased electrical impedance, and a steady increase in wall thickness with increasing BMI (Supplementary Figure 3e, 3h).

### Ethnicity

Across the two cohorts, 141 of 189 (74.6%) of non-white participants had T2DM, compared to white participants of which only 1640 of 3373 (48.6%) had T2DM. In both white and non-white subgroups, those with T2DM showed a significant increase in ventricular rate and an earlier T wave offset (Supplementary Table 6). A significant QRS shortening, QTc prolongation, T wave amplitude flattening, Sokolow-Lyon score reduction, lower end-diastolic, end-systolic, and stroke volumes, and thinner left ventricular wall were only observed in white participants. In contrast, no significant changes were observed in the non-white subgroup for these biomarkers.

### Association of HbA1c with ECG and CMR-derived biomarkers

In individuals with T2DM, we found a statistically significant association of higher levels of HbA1c with both a higher ventricular rate and an earlier T wave offset in all models (Supplementary Table 4). As noted earlier, heart rate and T wave offset are strongly correlated, so this consistent association is expected. Higher levels of HbA1c were also associated with a lower T wave amplitude in lead V3 and a lower stroke volume in all but the fully adjusted model. This suggests that, in addition to their association with binary T2DM status, variations in ventricular rate and T wave also have a significant association with changes in the glycaemic spectrum.

## Discussion

To our knowledge, this is the first large-scale study of multi-modal cardiac biomarkers in subjects with T2DM and no prior diagnosis of CVD. We demonstrated that T2DM is independently associated with significant, concurrent differences in ECG and CMR-derived biomarkers including a higher resting ventricular rate, prolonged QTc interval, reduced T wave amplitude, thicker left ventricular wall and lower stroke volume. These differences may reflect diabetes-induced pathophysiological changes, which we discuss below.

### Type 2 diabetes is associated with a higher resting heart rate, longer QTc interval and reduced T wave amplitude

T2DM was associated with a higher resting heart rate. High heart rates have been associated to hypertension and metabolic conditions for many decades [23,24]. In T2DM subjects specifically, this is typically a consequence of cardiac autonomic neuropathy (CAN) [7]. CAN causes heart rate to increase in the early stages of disease and gradually decrease back to normal, but with reduced variability and an increased likelihood of arrhythmias [25]. This may explain why heart rates in our T2DM cohort lie mostly within the normal clinical range of 60-100 beats per minute. Most subjects with T2DM were on anti-hypertensive (n=1003, 56.3%) and cholesterol-lowering medication (n=1287, 72.3%), likely due to concomitant hypertension and hyperlipidaemia, or as a preventative measure for CVD [2]. UK Biobank medication categories considered here are likely to include statins and beta-blockers, which are known to reduce heart rate and may have a disproportionate effect in the T2DM cohort [26,27]. Additionally, we found a significant association between HbA1c and resting heart rate (Supplementary Table 4). This supports evidence linking dysregulations in glucose metabolism with increased sympathetic nervous system activity and severity of CAN [28–30].

T2DM was also associated with a longer QTc interval. QTc prolongation is a well-established risk factor for cardiac arrhythmias and is correlated with a higher CAN score in patients with T2DM [11]. In our T2DM cohort, 113 women and 159 men (n=272 of 1781 [15%]) met sex- and age-specific criteria for clinical QTc prolongation [31]. Diabetic cardiomyocytes have a local renin-angiotensin system; when activated, increases in angiotensin II causes a reduction of transient outward potassium current Ito, extending the action potential repolarisation phase and thus prolonging the QT interval [5,32]. In addition, high glucose decreases hERG channel expression, which in turn modulates the delayed rectifier potassium current IKr, another factor responsible for QT prolongation in diabetes [33,34]. Hyperglycaemia has also been linked with a longer QT in clinical studies of type 1 diabetes [35,36]. Our results, however, did not show this association with HbA1c; mechanisms in T2DM may differ.

T2DM subjects exhibited lower T wave amplitudes in leads V3 and aVL. BMI was included both as a matching variable and as a model covariate, to minimise the effect of body composition on electrical impedance and subsequent reductions in ECG amplitude measurements. Thus, our results suggest a plausible direct effect of T2DM on T wave amplitude. Several studies have already demonstrated the presence of T wave abnormalities in T2DM subjects using the Minnesota coding system, indicating inverted T waves [12,37]. Hyperinsulinemic hypoglycaemia, which is common in T2DM, may reduce T wave amplitude [38]. A recent ECG study showed that T wave amplitudes were reduced in individuals with T2DM [39]. However, some subjects had a previous history of myocardial infarction, which itself is likely to affect T wave morphology. A reduced T wave amplitude has also been associated with hypokalaemia [40]. In T2DM, hypokalaemia and impaired insulin secretion have a bidirectional link [41]. Thus, the lower T wave amplitudes observed may reflect underlying changes in potassium, insulin or glucose levels, or the presence of silent ischemia.

### Type 2 diabetes is associated with increased left ventricular wall thickness and reduced stroke volume

In the T2DM cohort, the increase in heart rate (HR) was accompanied by a significant decrease in stroke volume (SV), while no change was observed in cardiac output (CO). Results suggest that the reduction in stroke volume in our T2DM cohort is most likely linked to a decreased end-diastolic volume. These results corroborate findings from a previous study examining CMR differences in a smaller cohort of 143 subjects with type 1 and 2 diabetes [8]. We also found that T2DM was associated with a thicker left ventricular wall, independently of blood pressure. This is consistent with previous studies that have demonstrated that patients with diabetes, hypertension, or both, tend to develop a thicker myocardial wall [42–44]. In later stages of disease, left ventricular hypertrophy is common in T2DM patients due to increased myocardial steatosis leading to hypertrophic signalling and concentric remodelling [45]. Coupled with a normal ejection fraction (>50%), both a lower stroke volume and thicker left ventricular wall can be indicators of heart failure with preserved ejection fraction (HFpEF), a frequent and serious complication of T2DM [46,47]. The changes observed may only represent early stages of adverse remodelling; measurements of left ventricular end-diastolic volume in our T2DM cohort fell within the normal sex-specific reference ranges that have been established for healthy adults without diabetes in the UK Biobank [20]. In the T2DM cohort, median wall thickness (6.1 mm) remained well below the clinical threshold for left ventricular hypertrophy (end-diastolic maximal wall thickness ≥ 15.0 mm) [48], and the absolute change compared to the control cohort was subtle (+0.2 mm).

Unexpectedly, the Sokolow-Lyon index was lower in the T2DM cohort, suggesting lower R and S wave amplitudes. The discordance between this index and left ventricular hypertrophy as assessed by CMR is interesting but plausible. In obese patients, QRS voltages are artificially reduced due to increased body fat causing electrical impedance [15]. Despite having similar BMI and left ventricular mass, subjects in different cohorts may have varying chest wall shapes, or additional myocardial fat deposition contributing to non-electrically active left ventricular mass, which may drive the observed difference in Sokolow-Lyon index.

### Clinical outcomes: development of cardiovascular disease

When comparing subjects who did and did not develop future CVD in the T2DM cohort, no statistically significant differences were observed in blood pressure, cholesterol, HbA1c, or eGFR. These biomarkers are clinical risk factors used in the calculation of SCORE2-Diabetes, a 10-year T2DM-specific cardiovascular risk prediction score developed by a working group of the European Society of Cardiology [49]. Here, the lack of significant differences may be due to a shorter follow-up time and pathophysiological changes still too subtle to be reflected, or a small sample size (n=98 CVD cases).

Changes in ECG biomarkers between CVD cases and controls differed in the cohorts with and without T2DM (Supplementary Table 2). Surprisingly, we observed no significant changes in heart rate between cases and controls in both cohorts, despite the established association between a higher heart rate and adverse cardiovascular outcomes [50,51]. In the T2DM cohort, 668 subjects (38%) had a heart rate above 70 bpm, a threshold associated with a higher risk of cardiovascular events specifically in T2DM [52]. An increased QRS duration was the only ECG biomarker showing a significant association with the development of cardiovascular disease in the T2DM cohort. In the ACCORD trial, QRS duration was increased in patients with diabetes and incident heart failure (HF) compared to those without HF [53]. Additionally, a longer QRS complex was established as an independent predictor of cardiovascular events in middle- to older-aged men and is associated with all-cause mortality in T2DM [54,55]. In contrast, in the control cohort, the Sokolow-Lyon index was the only ECG biomarker showing a significant association with the development of future CVD. This is in line with previous studies linking left ventricular hypertrophy with adverse cardiovascular events [56–60]. The hypothesis behind the hypertrophic CVD phenotype is strengthened by the strong independent association of increased left ventricular mass and wall thickness with development of CVD, which was present in both cohorts.

### Sex, age, body mass index and ethnicity-specific differences

Baseline biomarker values differed in males and females, which is expected [61–63]. The sex-specific subgroup analysis suggests that trends according to T2DM status were consistent across both sexes. The decrease in Sokolow-Lyon index observed across groups of increasing BMI is likely explained by increased electrical impedance due to a larger chest size.

In the subgroup of white ethnic background, trends in biomarker differences aligned with those of the full cohort. However, in non-white participants, we observed fewer significant differences. This lack of significance may be due partly to a smaller sample size (n=189). Indeed, we found a notable imbalance of participants in terms of ethnicity groups, with 1733 (97.3%) and 1640 (92.1%) of all participants with and without T2DM, respectively, being white. This is, however, reasonably representative of the national distribution for different ethnic groups in the UK population at the time of recruitment [64].

### Strengths and limitations

This study is the first to provide a concurrent analysis of ECG and CMR-derived biomarkers in T2DM specifically, as opposed to diabetes of unspecified or combined type, and no prior history of CVD. Thanks to the range of data available in the UK Biobank, we were able to characterise this large cohort in depth by capturing demographical and clinical data of the participants studied. These data were included as covariates in our regression analyses, accounting for key confounders and mediators. The UK Biobank’s robustly validated and systematic data recording protocol ensured that data collection bias was minimised [64]. Our study also has several limitations. Firstly, statistical significance does not necessarily equate to clinical relevance. Slight changes in certain biomarkers may not be much larger than natural population variations. This is particularly relevant to subgroup analyses involving smaller sample sizes, which may result in imprecise estimates due to random error. This applies specifically to groups with CVD outcomes (n=98) and non-white ethnicity (n=289, of which n=48 without T2DM). Regardless of sample size, we strived to interpret all statistically significant results cautiously and within the context of existing knowledge established by previously published research. Secondly, the lack of precision on compounds present in broad medication categories hampers our ability to disentangle direct effects of T2DM from the effect of specific medications. Another limitation is the lack of widespread UK Biobank linkage to primary care records. As of 2024, only 45% of the UK Biobank cohort was linked to these records for general research purposes [65]. This directly impacts cohort size and composition, especially for chronic conditions like diabetes which tend to be diagnosed and recorded within a primary care setting as opposed to hospital admissions. Linkage for the entire UK Biobank cohort would increase statistical power and robustness of future population studies. We believe that matching participants by BMI was adequate for previously stated reasons, however we acknowledge that waist-to-hip ratio is a better indicator of general health and mortality [66]. Finally, we recognise that including multiple distinct conditions within the broad umbrella of CVD may dissolve opposing trends in certain markers. This could be tackled by focusing on a single condition, albeit with small samples sizes.

## Conclusion

Our study examined multi-modal cardiac biomarkers of a large cohort of individuals with type 2 diabetes, a well-established high-risk factor for the development of cardiovascular disease. Despite a lack of clinical diagnosis of cardiovascular disease in the subjects studied, we found that type 2 diabetes was associated with an increased ventricular rate and an altered ECG including a prolonged QTc and flatter T wave, compared to control subjects matched by sex, age and BMI. Those ECG changes were accompanied by a decrease in stroke volume and thicker ventricular wall, characterising subclinical cardiovascular changes in diabetes. Our results provide further evidence supporting current clinical knowledge and hypotheses surrounding diabetes-induced pathophysiological alterations in the heart. Subject to further validation, our findings support the need for early screening to identify subclinical cardiac abnormalities in patients with type 2 diabetes and ultimately reduce risk of cardiovascular disease.

## Acknowledgements

The authors acknowledge and thank the UK Biobank participants whose data form the basis of this study. We also thank Hannah J. Smith and Associate Professor Jemma Hopewell for their insights on conducting studies with the UK Biobank.

Computational resources were supported by PRACE-ICEI projects icp013 and icp019, the EPSRC project CompBioMed X (EP/X019446/1) and the CompBioMed 2 Centre of Excellence in Computational Biomedicine (European Commission Horizon 2020 research and innovation programme, grant agreements No. 675451 and No. 823712, respectively), the Polaris machine at the Argonne Leadership Computing Facility (ALCF), Argonne National Laboratory, United States of America. The access to Polaris was awarded by the U.S. Department of Energy’s (DOE) Innovative and Novel Computational Impact on Theory and Experiment (INCITE) Program. The ACLF is supported by the Office of Science of the U.S. DOE under Contract No. DE-AC02-06CH11357.

## Data availability

The data underlying this article were provided by the UK Biobank upon application. Access to UK Biobank data for research purposes can be obtained upon application (https://www.ukbiobank.ac.uk/).

## References

1. Sun H, Saeedi P, Karuranga S et al. IDF Diabetes Atlas: Global, regional and country-level diabetes prevalence estimates for 2021 and projections for 2045. Diabetes Res Clin Pract 2022;183:109119.

2. Marx N, Federici M, Schütt K et al. 2023 ESC Guidelines for the management of cardiovascular disease in patients with diabetes: Developed by the task force on the management of cardiovascular disease in patients with diabetes of the European Society of Cardiology (ESC). Eur Heart J 2023;44:4043–140.

3. World Health Organisation. Cardiovascular diseases. https://www.who.int/health-topics/cardiovascular-diseases.

4. Gallego M, Zayas-Arrabal J, Alquiza A et al. Electrical Features of the Diabetic Myocardium. Arrhythmic and Cardiovascular Safety Considerations in Diabetes. Front Pharmacol 2021;12, DOI: 10.3389/fphar.2021.687256.

5. Ozturk N, Uslu S, Ozdemir S. Diabetes-induced changes in cardiac voltage-gated ion channels. World J Diabetes 2021;12:1–18.

6. Cheng Y, Wang Y, Yin R et al. Central role of cardiac fibroblasts in myocardial fibrosis of diabetic cardiomyopathy. Front Endocrinol (Lausanne) 2023;14, DOI: 10.3389/fendo.2023.1162754.

7. Vinik AI, Ziegler D. Diabetic Cardiovascular Autonomic Neuropathy. Circulation 2007;115:387–97.

8. Jensen MT, Fung K, Aung N et al. Changes in Cardiac Morphology and Function in Individuals With Diabetes Mellitus. Circ Cardiovasc Imaging 2019;12, DOI: 10.1161/CIRCIMAGING.119.009476.

9. Stern S, Sclarowsky S. The ECG in Diabetes Mellitus. Circulation 2009;120:1633–6.

10. Tokatli A, Kiliçaslan F, Alis M et al. Prolonged Tp-e Interval, Tp-e/QT Ratio and Tp-e/QTc Ratio in Patients with Type 2 Diabetes Mellitus. Endocrinology and Metabolism 2016;31:105.

11. Pappachan JM, Sebastian J, Bino BC et al. Cardiac autonomic neuropathy in diabetes mellitus: prevalence, risk factors and utility of corrected QT interval in the ECG for its diagnosis. Postgrad Med J 2008;84:205–10.

12. Mould SJ, Soliman EZ, Bertoni AG et al. Association of T-wave abnormalities with major cardiovascular events in diabetes: the ACCORD trial. Diabetologia 2021;64:504–11.

13. Petersen SE, Matthews PM, Francis JM et al. UK Biobank’s cardiovascular magnetic resonance protocol. Journal of Cardiovascular Magnetic Resonance 2016;18:8.

14. Eastwood S V, Mathur R, Atkinson M et al. Algorithms for the Capture and Adjudication of Prevalent and Incident Diabetes in UK Biobank. PLoS One 2016;11:e0162388.

15. Rider OJ, Ntusi N, Bull SC et al. Improvements in ECG accuracy for diagnosis of left ventricular hypertrophy in obesity. Heart 2016;102:1566–72.

16. Spiel C, Lapka D, Gradinger P et al. A Euclidean distance-based matching procedure for nonrandomized comparison studies. Eur Psychol 2008;13:180–7.

17. Levey AS, Stevens LA, Schmid CH et al. A New Equation to Estimate Glomerular Filtration Rate. Ann Intern Med 2009;150:604.

18. Locatelli F, Aljama P, Barany P et al. Foreword. Nephrology Dialysis Transplantation 2004;19:ii1–ii1.

19. Sokolow M, Lyon TP. The ventricular complex in left ventricular hypertrophy as obtained by unipolar precordial and limb leads. Am Heart J 1949;37:161–86.

20. Petersen SE, Aung N, Sanghvi MM et al. Reference ranges for cardiac structure and function using cardiovascular magnetic resonance (CMR) in Caucasians from the UK Biobank population cohort. Journal of Cardiovascular Magnetic Resonance 2016;19:18.

21. Bai W, Suzuki H, Huang J et al. A population-based phenome-wide association study of cardiac and aortic structure and function. Nat Med 2020;26:1654–62.

22. Johnson NP. Metformin use in women with polycystic ovary syndrome. Ann Transl Med 2014;2:56.

23. Ewing DJ, Campbell IW, Clarke BF. Heart rate changes in diabetes mellitus. The Lancet 1981;317.

24. Palatini P, Casiglia E, Pauletto P et al. Relationship of Tachycardia With High Blood Pressure and Metabolic Abnormalities. Hypertension 1997;30:1267–73.

25. Vinik AI, Maser RE, Ziegler D. Autonomic imbalance: prophet of doom or scope for hope? Diabetic Medicine 2011;28:643–51.

26. Liu HT, Deng NH, Wu ZF et al. Statin’s role on blood pressure levels: Meta-analysis based on randomized controlled trials. The Journal of Clinical Hypertension 2023;25:238–50.

27. do Vale GT, Ceron CS, Gonzaga NA et al. Three Generations of β-blockers: History, Class Differences and Clinical Applicability. Curr Hypertens Rev 2019;15:22–31.

28. Yeap BB, Russo A, Fraser RJ et al. Hyperglycemia affects cardiovascular autonomic nerve function in normal subjects. Diabetes Care 1996;19:880–2.

29. Synowski SJ, Kop WJ, Warwick ZS et al. Effects of glucose ingestion on autonomic and cardiovascular measures during rest and mental challenge. J Psychosom Res 2013;74:149–54.

30. Lai Y-R, Huang C-C, Chiu W-C et al. HbA1C Variability Is Strongly Associated With the Severity of Cardiovascular Autonomic Neuropathy in Patients With Type 2 Diabetes After Longer Diabetes Duration. Front Neurosci 2019;13, DOI: 10.3389/fnins.2019.00458.

31. Rautaharju PM, Mason JW, Akiyama T. New age- and sex-specific criteria for QT prolongation based on rate correction formulas that minimize bias at the upper normal limits. Int J Cardiol 2014;174:535–40.

32. Jermendy G, Koltai MZ, Pogátsa G. QT interval prolongation in type 2 (non-insulin-dependent) diabetic patients with cardiac autonomic neuropathy. Acta Diabetol Lat 1990;27:295–301.

33. Shi Y-Q, Yan M, Liu L-R et al. High Glucose Represses hERG K+ Channel Expression through Trafficking Inhibition. Cellular Physiology and Biochemistry 2015;37:284–96.

34. Zhang Y, Xiao J, Lin H et al. Ionic Mechanisms Underlying Abnormal QT Prolongation and the Associated Arrhythmias in Diabetic Rabbits: A Role of Rapid Delayed Rectifier K+ Current. Cellular Physiology and Biochemistry 2007;19:225–38.

35. Gordin D, Forsblom C, Rönnback M et al. Acute hyperglycaemia disturbs cardiac repolarization in Type 1 diabetes. Diabetic Medicine 2008;25:101–5.

36. Stern K, Cho YH, Benitez-Aguirre P et al. QT interval, corrected for heart rate, is associated with HbA1c concentration and autonomic function in diabetes. Diabetic Medicine 2016;33:1415–21.

37. Soflaei Saffar S, Nazar E, Sahranavard T et al. Association of T-wave electrocardiogram changes and type 2 diabetes: a cross-sectional sub-analysis of the MASHAD cohort population using the Minnesota coding system. BMC Cardiovasc Disord 2024;24:48.

38. Laitinen T, Lyyra-Laitinen T, Huopio H et al. Electrocardiographic Alterations during Hyperinsulinemic Hypoglycemia in Healthy Subjects. Annals of Noninvasive Electrocardiology 2008;13:97–105.

39. Isaksen JL, Sivertsen CB, Jensen CZ et al. Electrocardiographic markers in patients with type 2 diabetes and the role of diabetes duration. J Electrocardiol 2024;84:129–36.

40. Diercks DB, Shumaik GM, Harrigan RA et al. Electrocardiographic manifestations: electrolyte abnormalities. J Emerg Med 2004;27:153–60.

41. Liamis G, Liberopoulos E, Barkas F et al. Diabetes mellitus and electrolyte disorders. World J Clin Cases 2014;2:488.

42. Devereux RB, Roman MJ, Paranicas M et al. Impact of Diabetes on Cardiac Structure and Function. Circulation 2000;101:2271–6.

43. Palmieri V, Bella JN, Arnett DK et al. Effect of Type 2 Diabetes Mellitus on Left Ventricular Geometry and Systolic Function in Hypertensive Subjects. Circulation 2001;103:102–7.

44. Eguchi K, Kario K, Hoshide S et al. Type 2 diabetes is associated with left ventricular concentric remodeling in hypertensive patients. Am J Hypertens 2005;18:23–9.

45. Levelt E, Mahmod M, Piechnik SK et al. Relationship Between Left Ventricular Structural and Metabolic Remodeling in Type 2 Diabetes. Diabetes 2016;65:44–52.

46. Aurigemma GP, Zile MR, Gaasch WH. Contractile Behavior of the Left Ventricle in Diastolic Heart Failure. Circulation 2006;113:296–304.

47. Paulus WJ, Tschöpe C. A Novel Paradigm for Heart Failure With Preserved Ejection Fraction. J Am Coll Cardiol 2013;62:263–71.

48. Authors/Task Force members, Elliott PM, Anastasakis A et al. 2014 ESC Guidelines on diagnosis and management of hypertrophic cardiomyopathy: The Task Force for the Diagnosis and Management of Hypertrophic Cardiomyopathy of the European Society of Cardiology (ESC). Eur Heart J 2014;35:2733–79.

49. SCORE2-Diabetes Working Group and the ESC Cardiovascular Risk Collaboration. SCORE2-Diabetes: 10-year cardiovascular risk estimation in type 2 diabetes in Europe. Eur Heart J 2023;44:2544–56.

50. Medalie JH, Kahn HA, Neufeld HN et al. Five-year myocardial infarction incidence—II. Association of single variables to age and birthplace. J Chronic Dis 1973;26:329–49.

51. Böhm M, Schumacher H, Teo KK et al. Resting heart rate and cardiovascular outcomes in diabetic and non-diabetic individuals at high cardiovascular risk analysis from the ONTARGET/TRANSCEND trials. Eur Heart J 2020;41:231–8.

52. Ikeda S, Shinohara K, Enzan N et al. A higher resting heart rate is associated with cardiovascular event risk in patients with type 2 diabetes mellitus without known cardiovascular disease. Hypertension Research 2023;46:1090–9.

53. Segar MW, Vaduganathan M, Patel K V. et al. Machine Learning to Predict the Risk of Incident Heart Failure Hospitalization Among Patients With Diabetes: The WATCH-DM Risk Score. Diabetes Care 2019;42:2298–306.

54. Chen X, Hansson P-O, Thunström E et al. Incremental changes in QRS duration as predictor for cardiovascular disease: a 21-year follow-up of a randomly selected general population. Sci Rep 2021;11:13652.

55. Singleton MJ, German C, Hari KJ et al. QRS duration is associated with all-cause mortality in type 2 diabetes: The diabetes heart study. J Electrocardiol 2020;58:150–4.

56. Levy D, Garrison RJ, Savage DD et al. Prognostic Implications of Echocardiographically Determined Left Ventricular Mass in the Framingham Heart Study. New England Journal of Medicine 1990;322:1561–6.

57. Cooper RS, Simmons BE, Castaner A et al. Left ventricular hypertrophy is associated with worse survival independent of ventricular function and number of coronary arteries severely narrowed. Am J Cardiol 1990;65:441–5.

58. Koren MJ, Devereux RB, Casale PN et al. Relation of Left Ventricular Mass and Geometry to Morbidity and Mortality in Uncomplicated Essential Hypertension. Ann Intern Med 1991;114:345–52.

59. Aronow WS, Ahn C, Kronzon I et al. Congestive heart failure, coronary events and atherothrombotic brain infarction in elderly blacks and whites with systemic hypertension and with and without echocardiographic and electrocardiographic evidence of left ventricular hypertrophy. Am J Cardiol 1991;67:295–9.

60. Verdecchia P, Schillaci G, Borgioni C et al. Prognostic value of left ventricular mass and geometry in systemic hypertension with left ventricular hypertrophy. Am J Cardiol 1996;78:197–202.

61. James AF, Choisy SCM, Hancox JC. Recent advances in understanding sex differences in cardiac repolarization. Prog Biophys Mol Biol 2007;94:265–319.

62. Macfarlane PW. The Influence of Age and Sex on the Electrocardiogram. 2018, 93–106.

63. St. Pierre SR, Peirlinck M, Kuhl E. Sex Matters: A Comprehensive Comparison of Female and Male Hearts. Front Physiol 2022;13, DOI: 10.3389/fphys.2022.831179.

64. Sudlow C, Gallacher J, Allen N et al. UK Biobank: An Open Access Resource for Identifying the Causes of a Wide Range of Complex Diseases of Middle and Old Age. PLoS Med 2015;12:e1001779.

65. Allen NE, Lacey B, Lawlor DA et al. Prospective study design and data analysis in UK Biobank. Sci Transl Med 2024;16, DOI: 10.1126/scitranslmed.adf4428.

66. Khan I, Chong M, Le A et al. Surrogate Adiposity Markers and Mortality. JAMA Netw Open 2023;6:e2334836.

