## Supplementary Figures for "Multi-modal characterisation of cardiac function and electrophysiology in type 2 diabetes: a UK Biobank cross-sectional study"

Figure 1. Number of incident cardiovascular diagnoses in the control and type 2 diabetes cohorts. Shaded areas in each bar represent cases with a diagnosis of one or more other conditions.

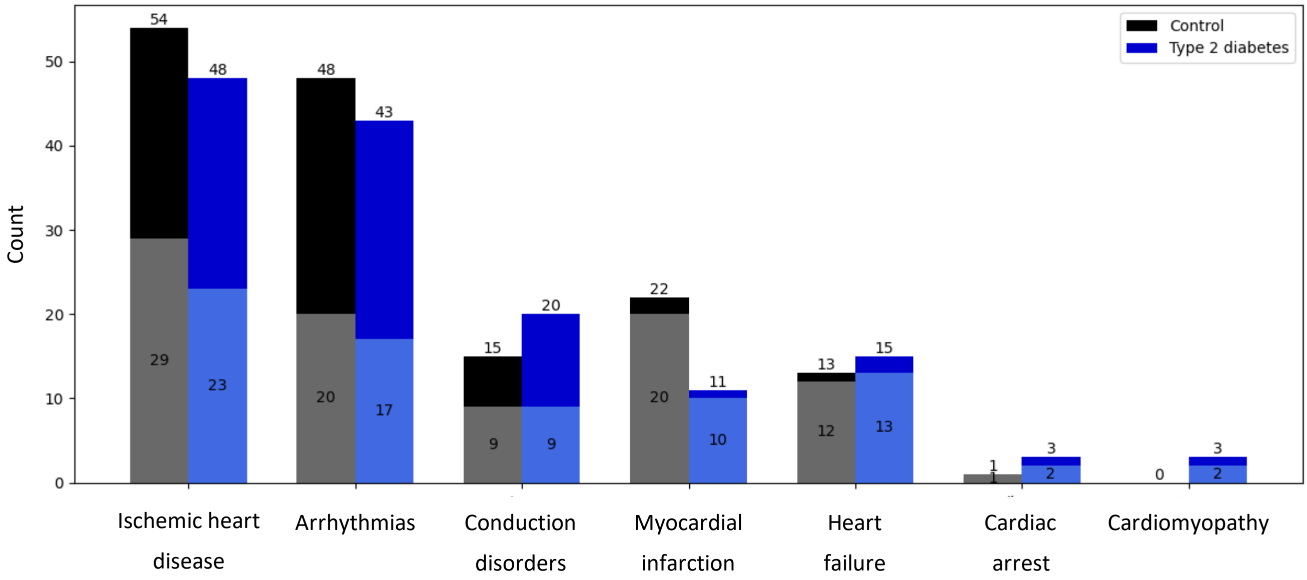

Figure 2. Matrix of Pearson correlation coefficients between potential model covariates. Coefficients were computed on 2620 cases; these cases contained no missing data among the features of interest. BMI: body mass index, HDL: high density lipoprotein, eGFR: estimated glomerular filtration rate.

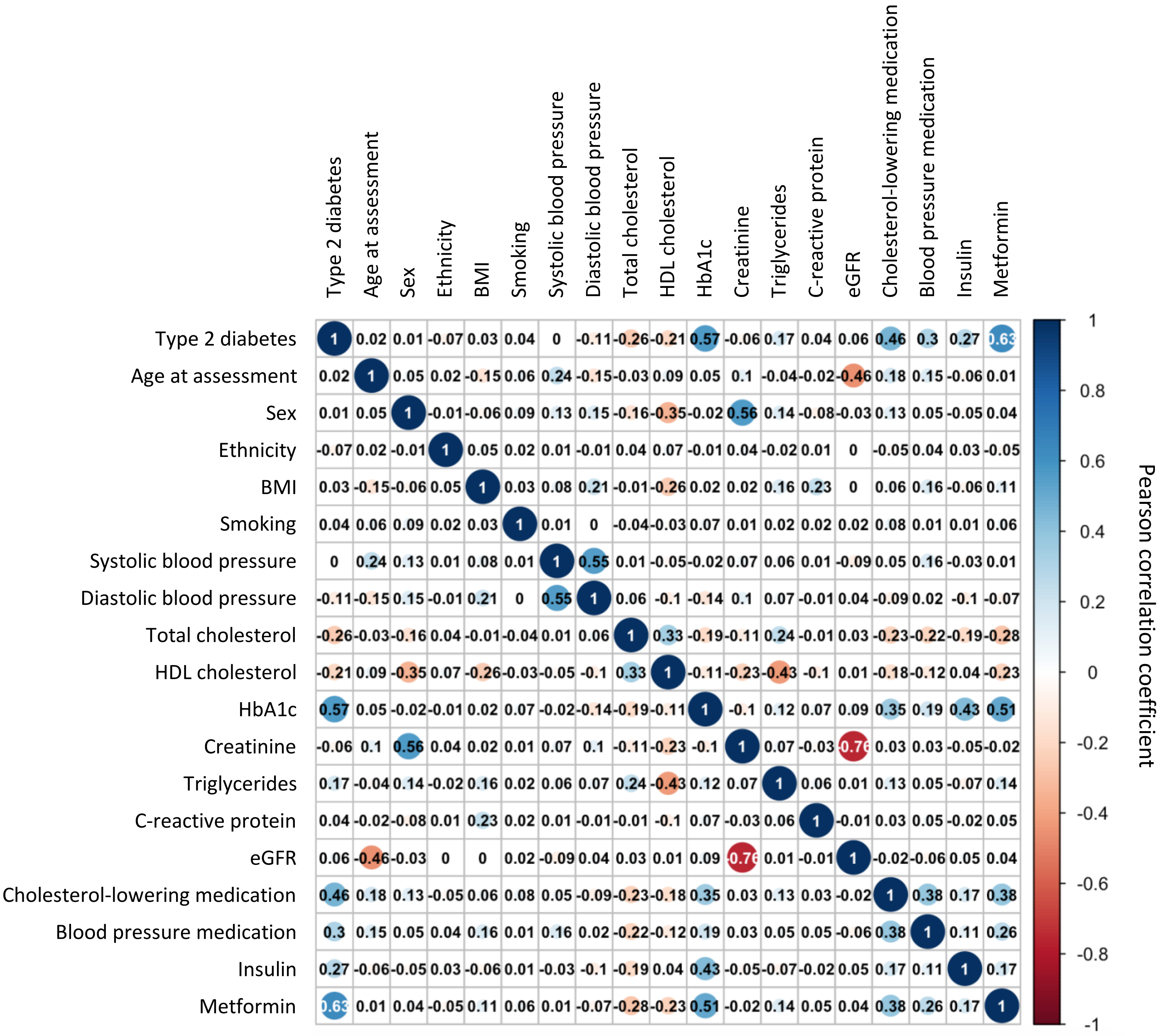

Figure 3. Distribution of sex-specific ECG and CMR-derived biomarkers stratified by age. LV: left ventricular, EF: ejection fraction. Cases with LVEF <20%, QTc interval >600ms and LV stroke volume >300ml are considered outliers and are not shown in the plots.

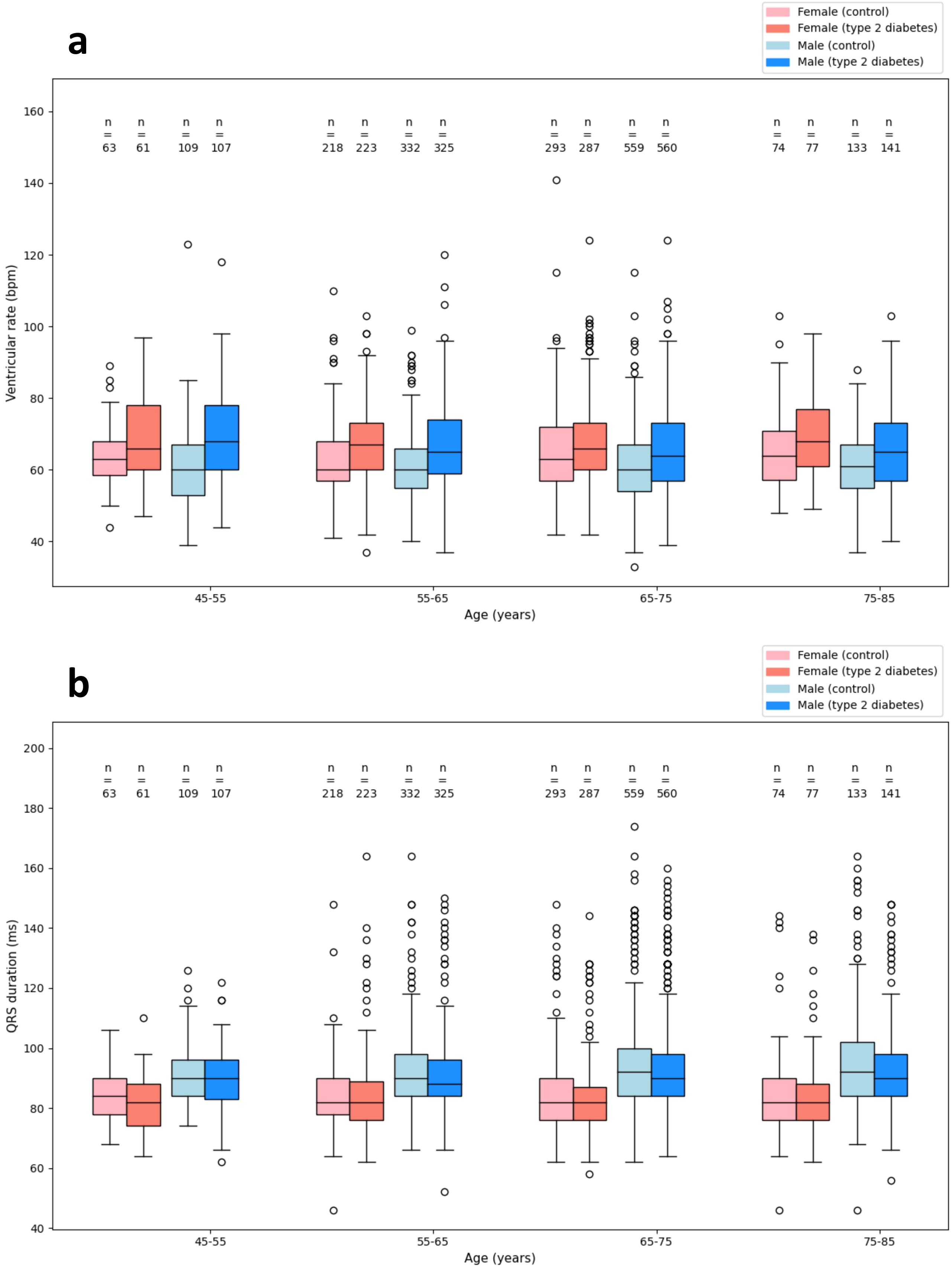

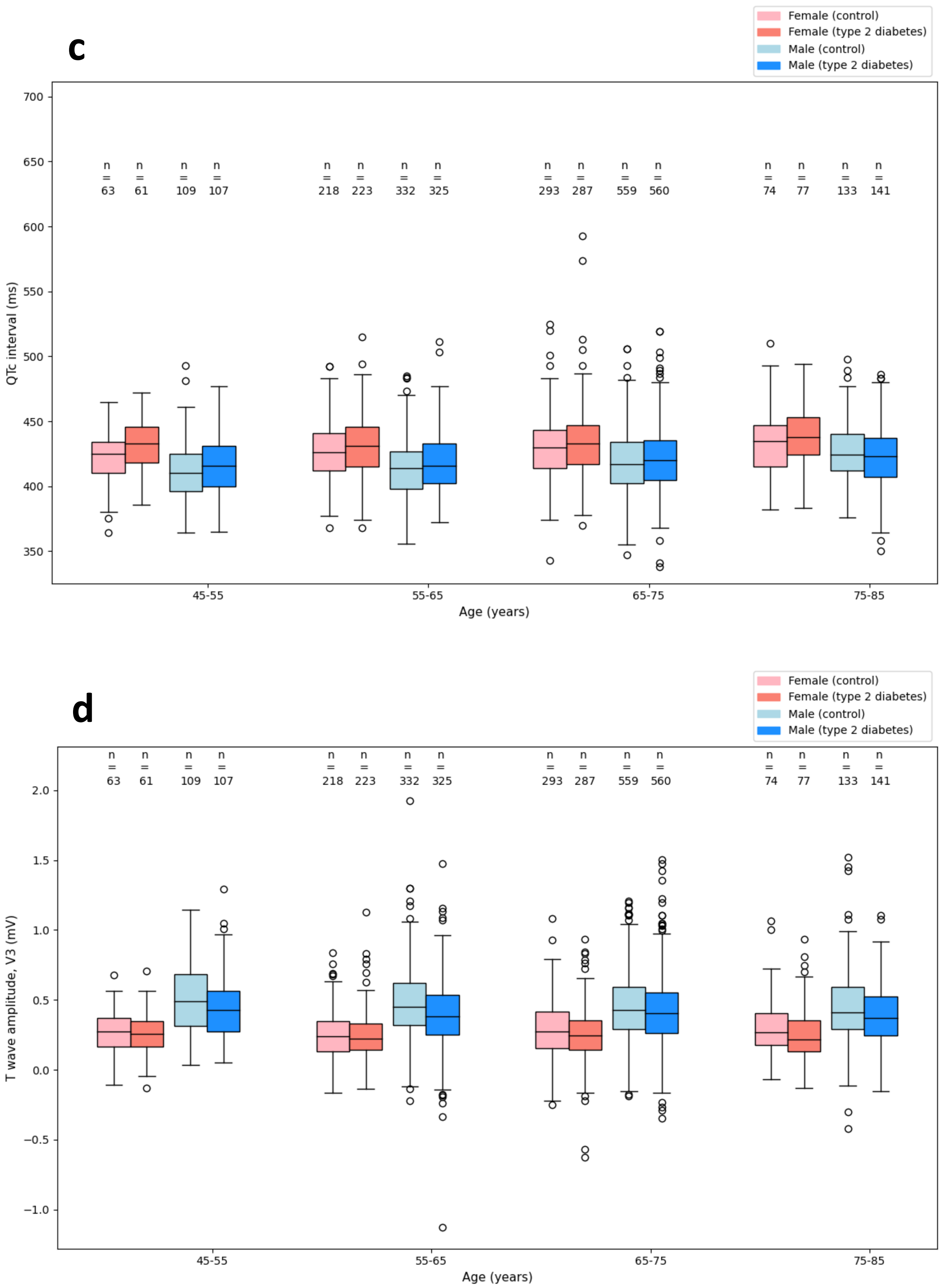
c
Female (control)
Female (type 2 diabetes)
Male (control)
Male (type 2 diabetes)
QTc interval (ms)
n = 63, n = 61, n = 109, n = 107
n = 218, n = 223, n = 332, n = 325
n = 293, n = 287, n = 559, n = 560
n = 74, n = 77, n = 133, n = 141
45-55
55-65
65-75
75-85
Age (years)
d
Female (control)
Female (type 2 diabetes)
Male (control)
Male (type 2 diabetes)
T wave amplitude, V3 (mV)
n = 63, n = 61, n = 109, n = 107
n = 218, n = 223, n = 332, n = 325
n = 293, n = 287, n = 559, n = 560
n = 74, n = 77, n = 133, n = 141
45-55
55-65
65-75
75-85
Age (years)

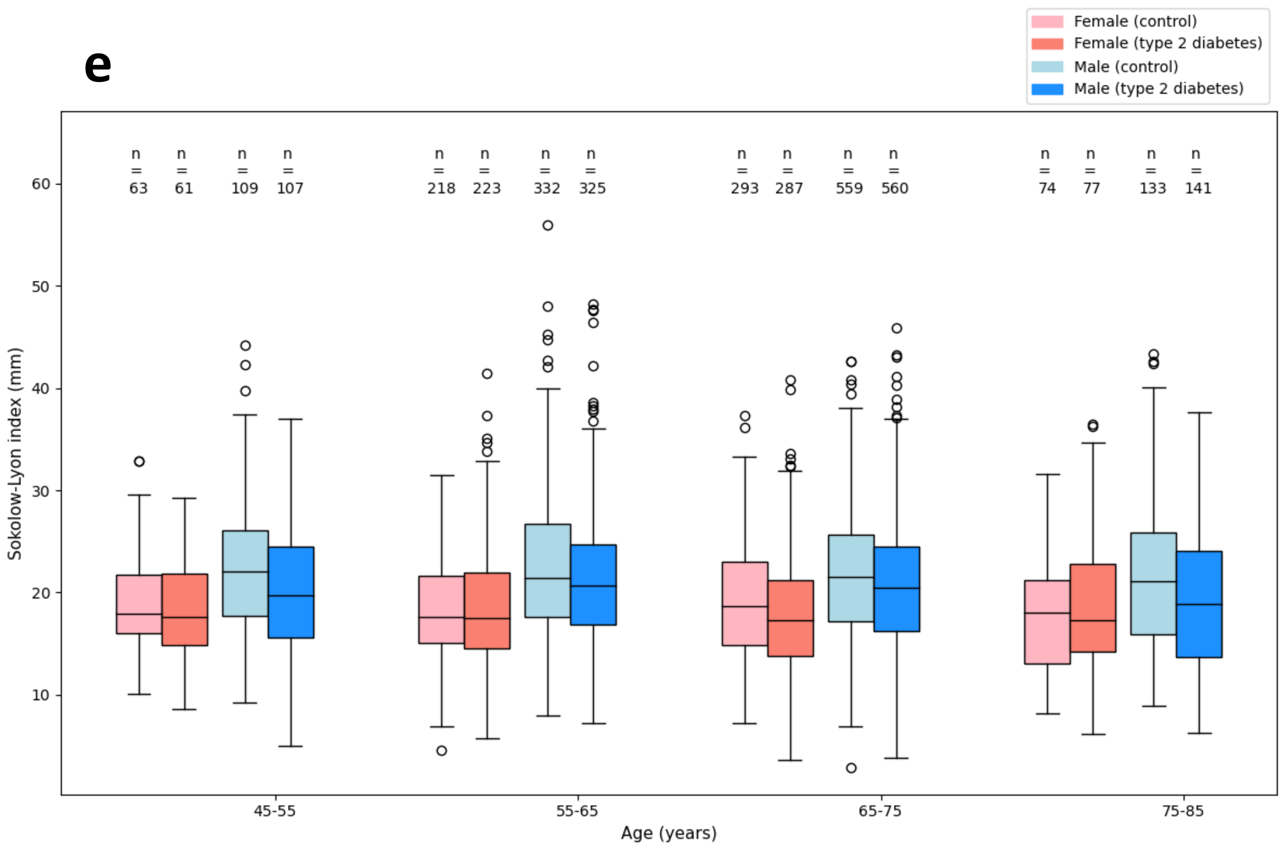
e
Female (control)
Female (type 2 diabetes)
Male (control)
Male (type 2 diabetes)
n = 63
n = 61
n = 109
n = 107
n = 218
n = 223
n = 332
n = 325
n = 293
n = 287
n = 559
n = 560
n = 74
n = 77
n = 133
n = 141
Sokolow-Lyon index (mm)
10
20
30
40
50
60
45-55
55-65
65-75
75-85
Age (years)

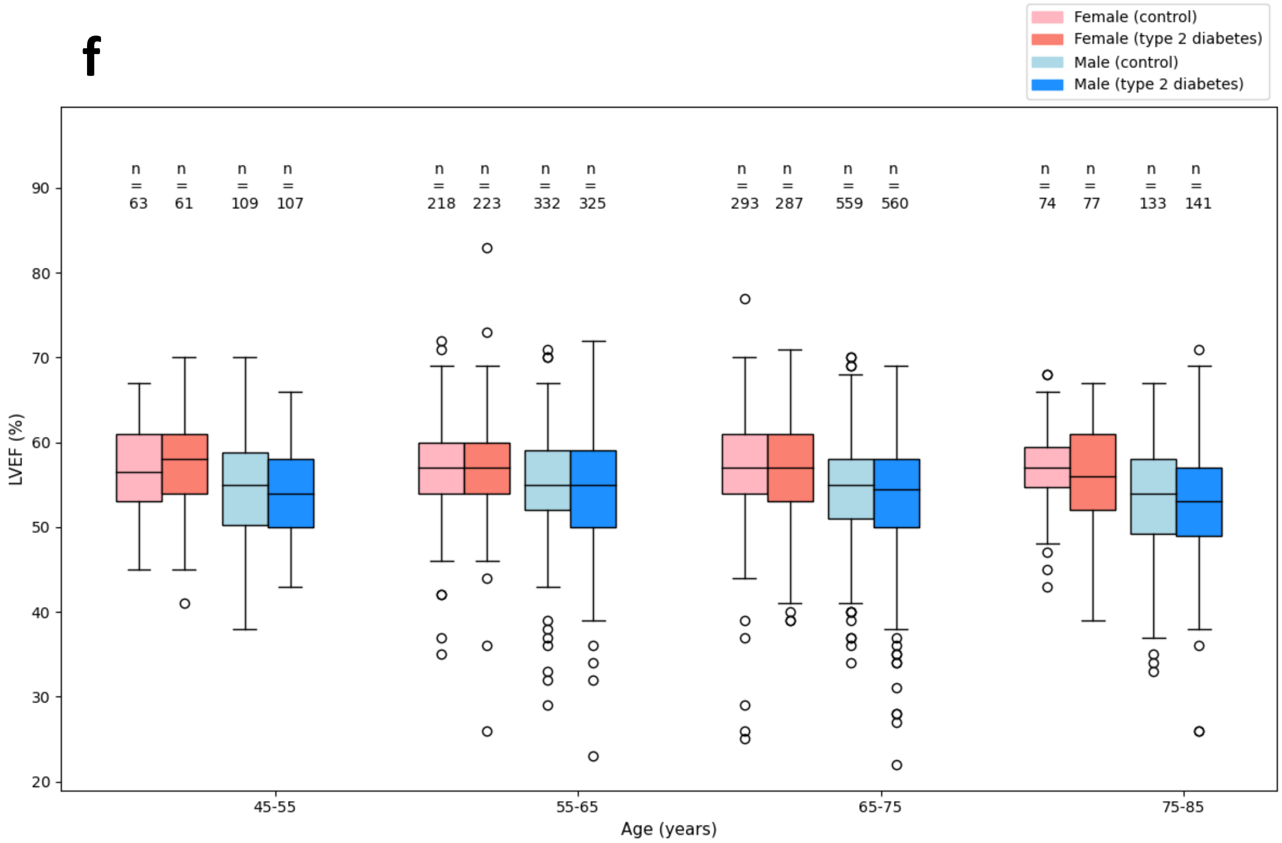
f
Female (control)
Female (type 2 diabetes)
Male (control)
Male (type 2 diabetes)
n = 63
n = 61
n = 109
n = 107
n = 218
n = 223
n = 332
n = 325
n = 293
n = 287
n = 559
n = 560
n = 74
n = 77
n = 133
n = 141
LVEF (%)
20
30
40
50
60
70
80
90
45-55
55-65
65-75
75-85
Age (years)

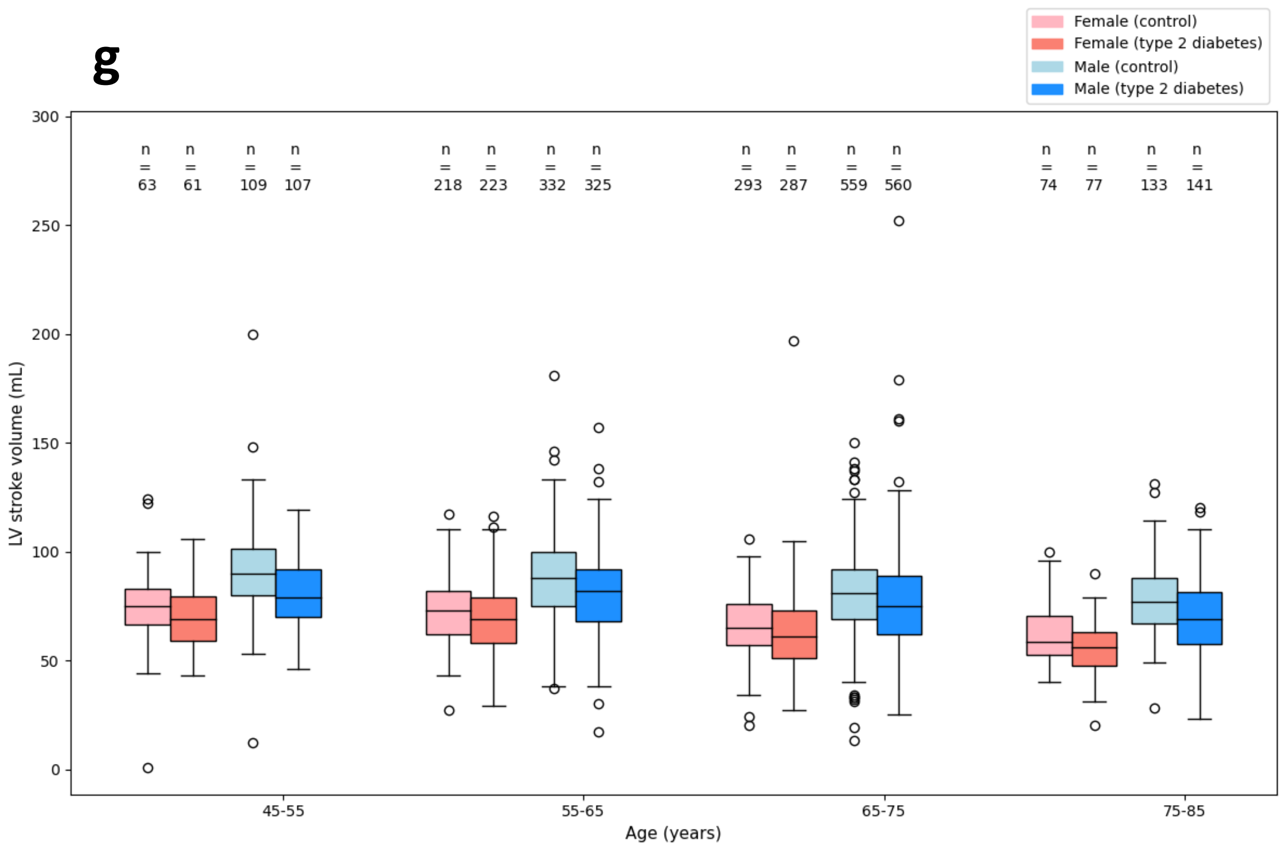
g
Female (control)
Female (type 2 diabetes)
Male (control)
Male (type 2 diabetes)
LV stroke volume (mL)
n = 63
n = 61
n = 109
n = 107
n = 218
n = 223
n = 332
n = 325
n = 293
n = 287
n = 559
n = 560
n = 74
n = 77
n = 133
n = 141
45-55
55-65
65-75
75-85
Age (years)

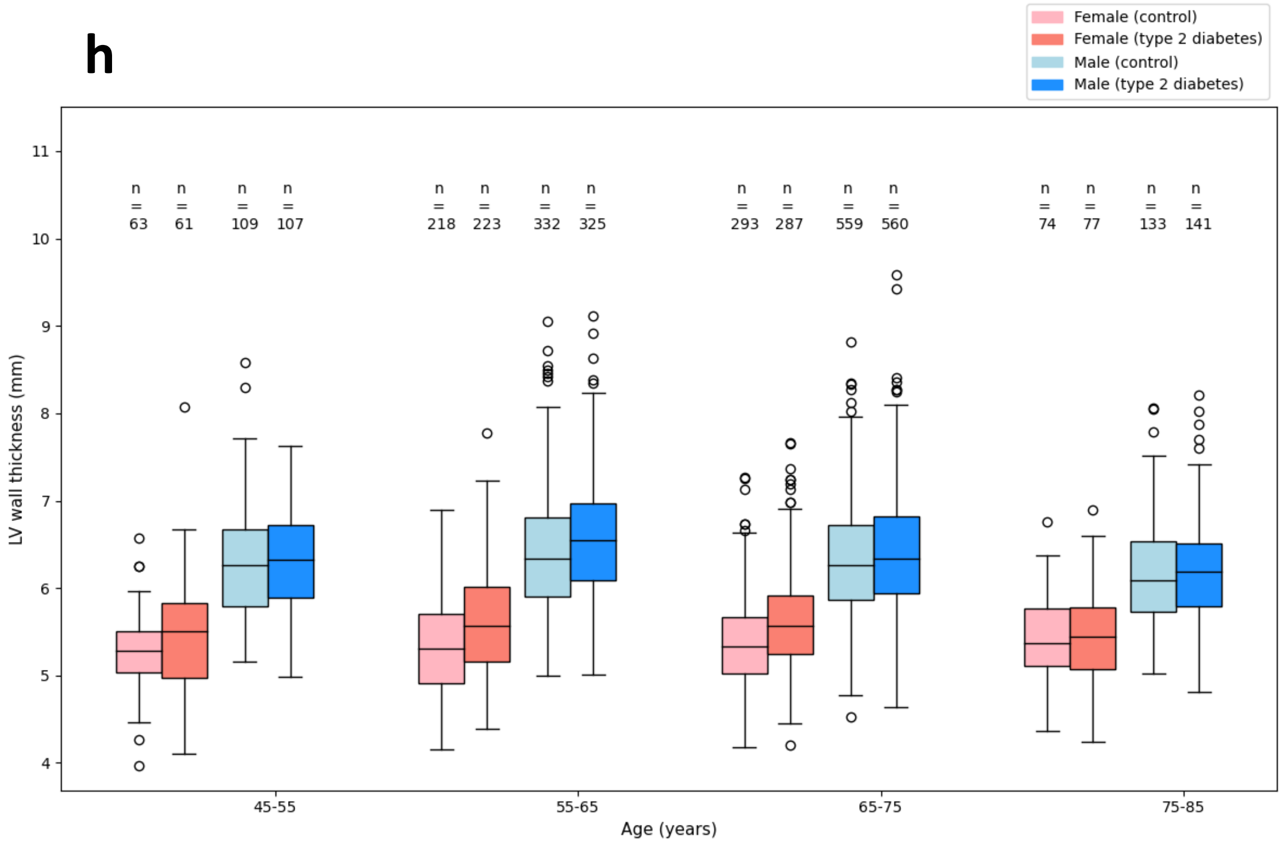
h
Female (control)
Female (type 2 diabetes)
Male (control)
Male (type 2 diabetes)
LV wall thickness (mm)
n = 63
n = 61
n = 109
n = 107
n = 218
n = 223
n = 332
n = 325
n = 293
n = 287
n = 559
n = 560
n = 74
n = 77
n = 133
n = 141
45-55
55-65
65-75
75-85
Age (years)

Figure 4. Distribution of sex-specific ECG and CMR-derived biomarkers stratified by body mass index. LV: left ventricular, EF: ejection fraction. Cases with LVEF <20%, QTc interval >600ms and LV stroke volume >300ml are considered outliers and are not shown in the plots.

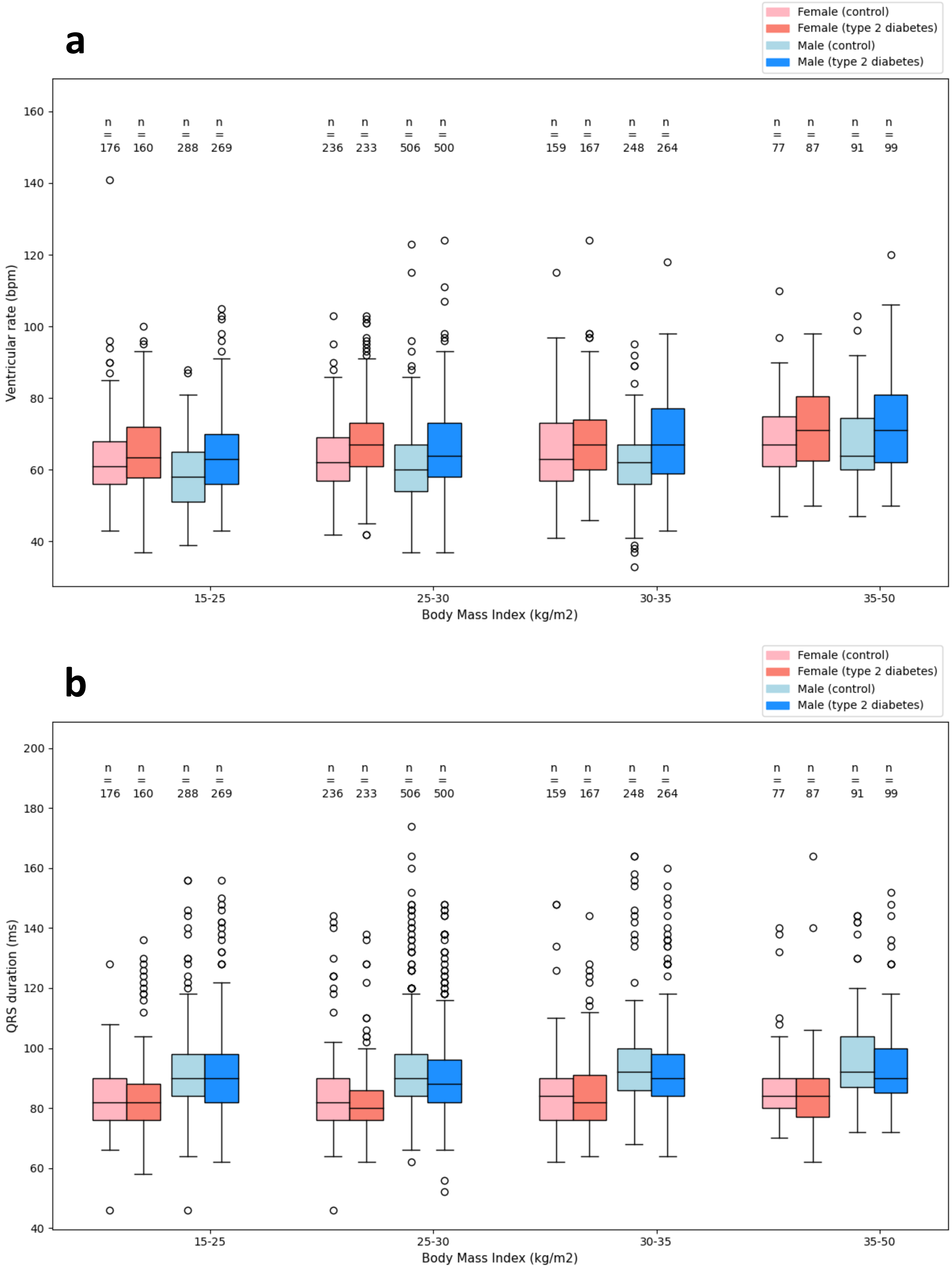

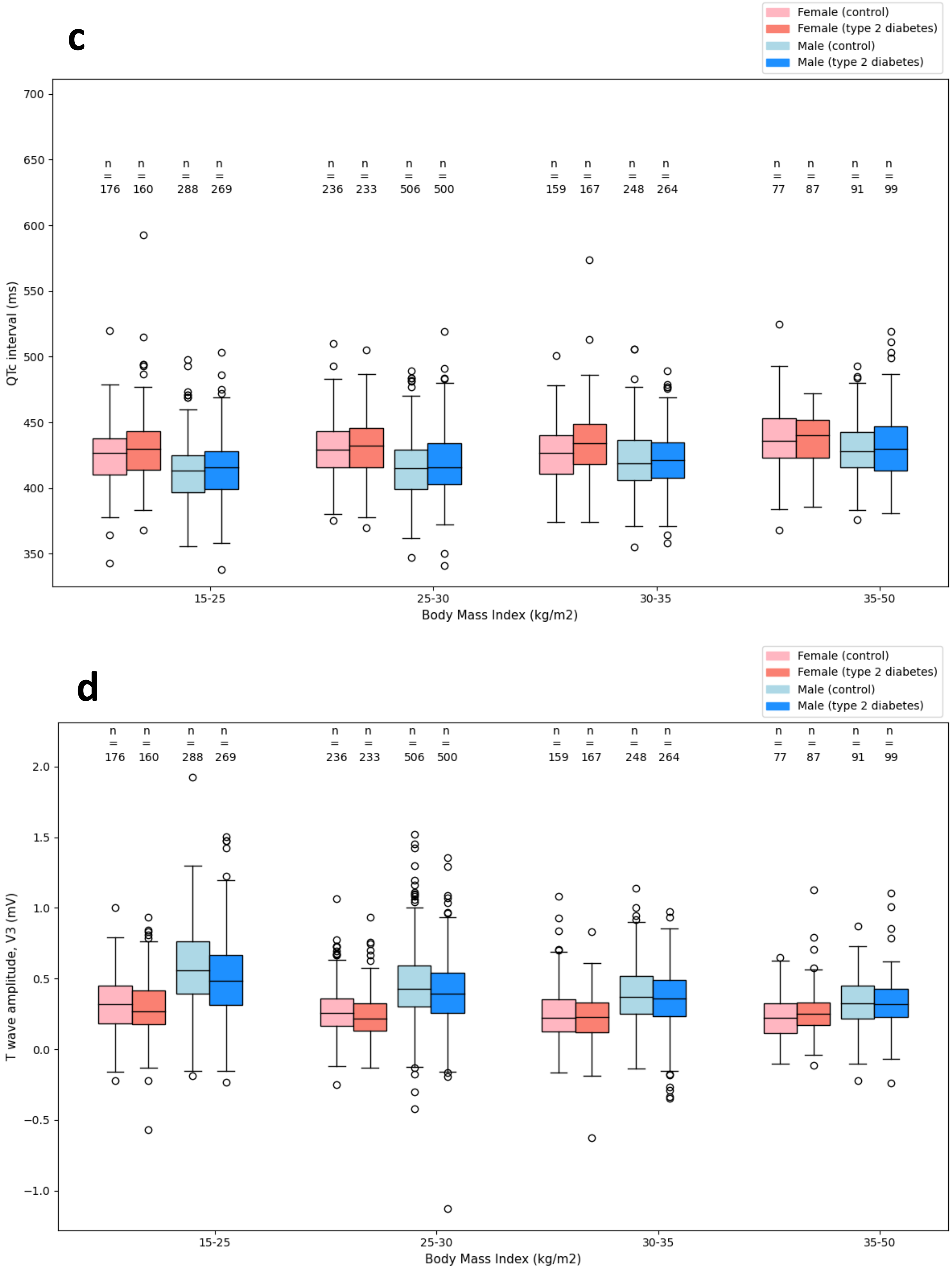
c
Female (control)
Female (type 2 diabetes)
Male (control)
Male (type 2 diabetes)
QTc interval (ms)
700
650
600
550
500
450
400
350
n = 176
n = 160
n = 288
n = 269
n = 236
n = 233
n = 506
n = 500
n = 159
n = 167
n = 248
n = 264
n = 77
n = 87
n = 91
n = 99
15-25
25-30
30-35
35-50
Body Mass Index (kg/m2)
d
Female (control)
Female (type 2 diabetes)
Male (control)
Male (type 2 diabetes)
T wave amplitude, V3 (mV)
2.0
1.5
1.0
0.5
0.0
−0.5
−1.0
n = 176
n = 160
n = 288
n = 269
n = 236
n = 233
n = 506
n = 500
n = 159
n = 167
n = 248
n = 264
n = 77
n = 87
n = 91
n = 99
15-25
25-30
30-35
35-50
Body Mass Index (kg/m2)

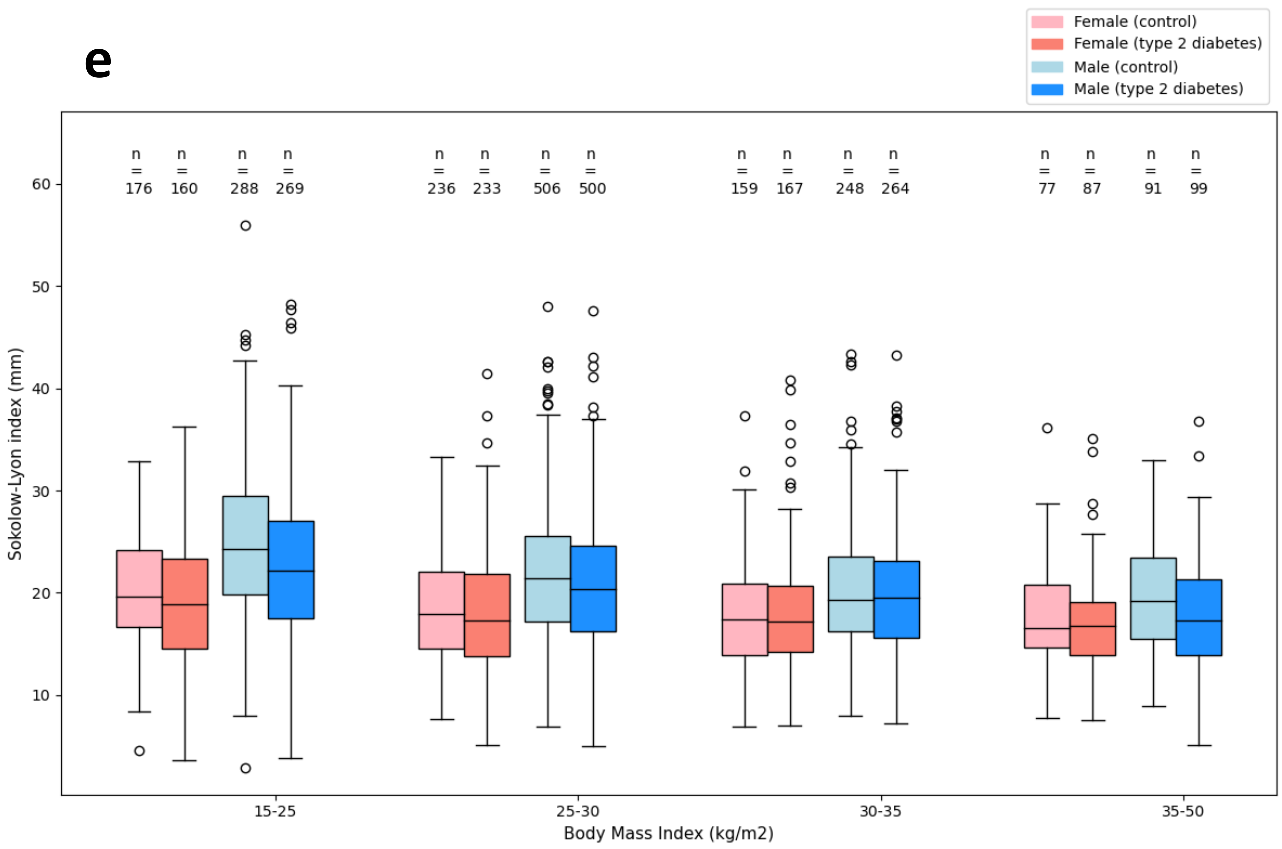
e
Female (control)
Female (type 2 diabetes)
Male (control)
Male (type 2 diabetes)
n = 176
n = 160
n = 288
n = 269
n = 236
n = 233
n = 506
n = 500
n = 159
n = 167
n = 248
n = 264
n = 77
n = 87
n = 91
n = 99
Sokolow-Lyon index (mm)
15-25
25-30
30-35
35-50
Body Mass Index (kg/m2)

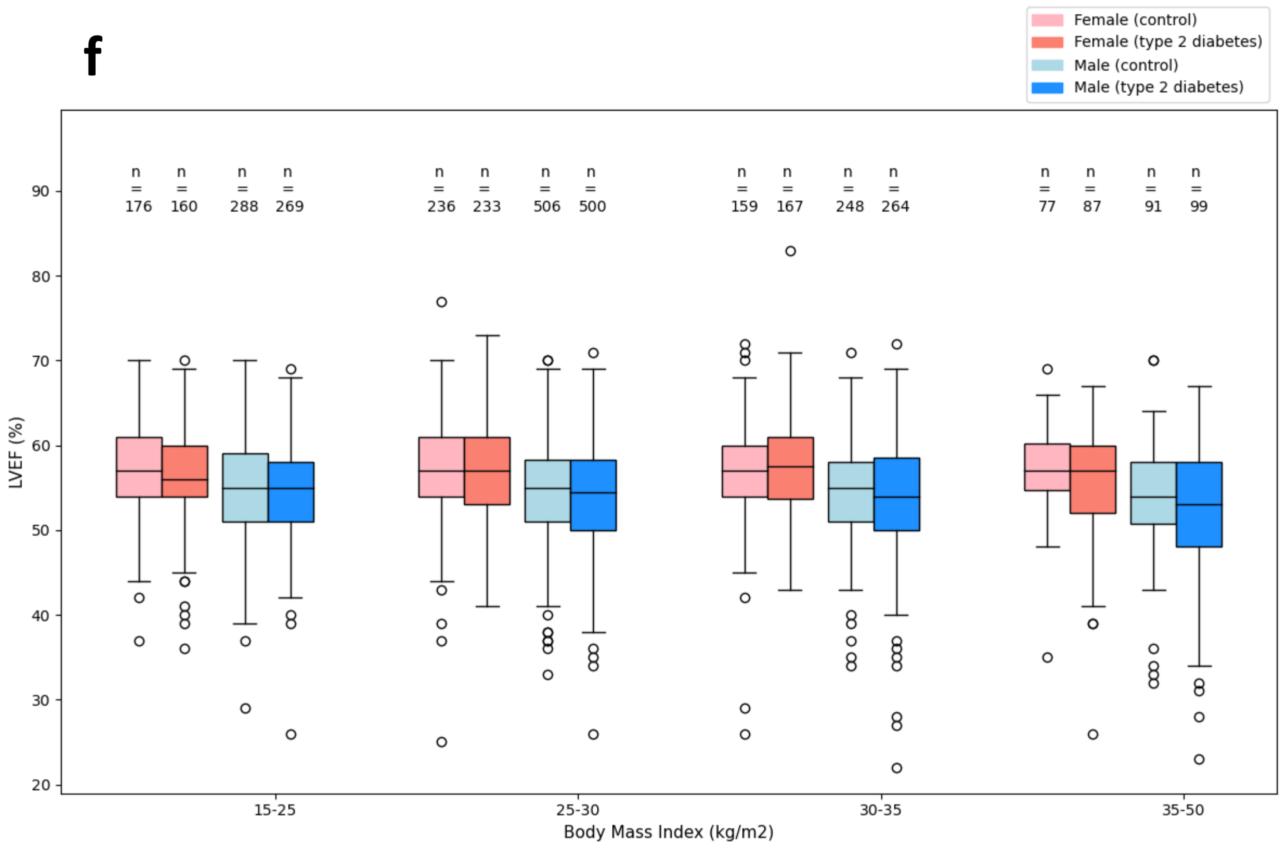
f
Female (control)
Female (type 2 diabetes)
Male (control)
Male (type 2 diabetes)
n = 176
n = 160
n = 288
n = 269
n = 236
n = 233
n = 506
n = 500
n = 159
n = 167
n = 248
n = 264
n = 77
n = 87
n = 91
n = 99
LVEF (%)
15-25
25-30
30-35
35-50
Body Mass Index (kg/m2)

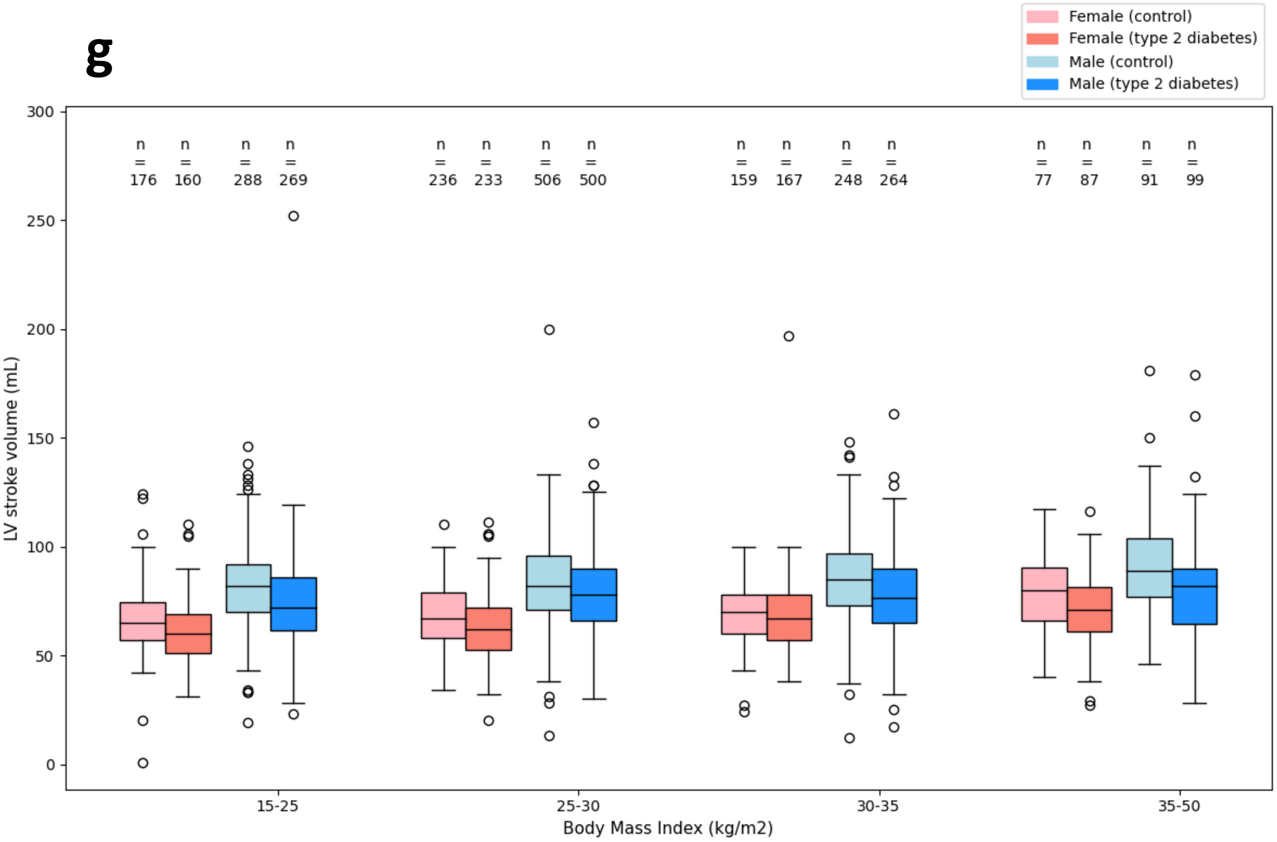
g
Female (control)
Female (type 2 diabetes)
Male (control)
Male (type 2 diabetes)
n = 176
n = 160
n = 288
n = 269
n = 236
n = 233
n = 506
n = 500
n = 159
n = 167
n = 248
n = 264
n = 77
n = 87
n = 91
n = 99
LV stroke volume (mL)
0
50
100
150
200
250
300
15-25
25-30
30-35
35-50
Body Mass Index (kg/m2)

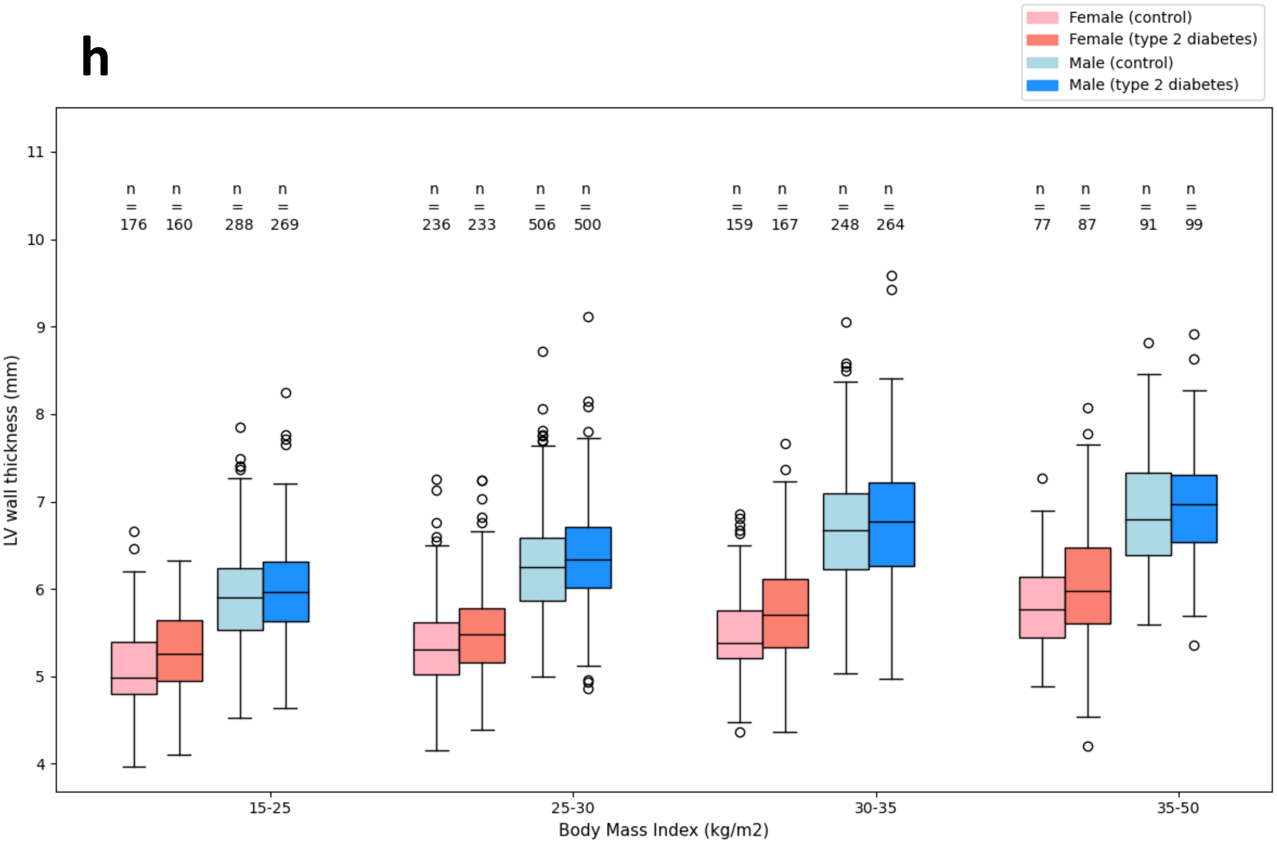
h
Female (control)
Female (type 2 diabetes)
Male (control)
Male (type 2 diabetes)
n = 176
n = 160
n = 288
n = 269
n = 236
n = 233
n = 506
n = 500
n = 159
n = 167
n = 248
n = 264
n = 77
n = 87
n = 91
n = 99
LV wall thickness (mm)
4
5
6
7
8
9
10
11
15-25
25-30
30-35
35-50
Body Mass Index (kg/m2)
