## Supplementary Tables for "Multi-modal characterisation of cardiac function and electrophysiology in type 2 diabetes: a UK Biobank cross-sectional study"

Table 1. Selected ICD-9 and ICD-10 codes for type 2 diabetes and cardiovascular disease used to define the study cohorts. UK Biobank fields corresponding to the first reported date of ICD-10 code are indicated in the right column. No equivalent fields exist for ICD-9 codes.

| **ICD-X code** | | **Description** | **UK Biobank data field for "Date [ICD-10 code] first reported"** |
|---|---|---|---|
| **Type 2 diabetes** | | | |
| ICD-9 | 250.x0 | diabetes mellitus, type II or unspecified type, not stated as uncontrolled | - |
| | 250.x2 | diabetes mellitus, type II or unspecified type, uncontrolled | - |
| ICD-10 | E11 | type 2 diabetes mellitus | 130708 |
| | E14 | unspecified diabetes mellitus | 130714 |
| **Cardiovascular disease** | | | |
| ICD-9 | 410 | Acute myocardial infarction | - |
| | 411 | Other acute and subacute forms of ischaemic heart disease | - |
| | 412 | Old myocardial infarction | - |
| | 413 | Angina pectoris | - |
| | 414 | Other forms of chronic ischaemic heart disease | - |
| | 425 | Cardiomyopathy | - |
| | 426 | Conduction disorders | - |
| | 427 | Cardiac dysrhythmias | - |
| | 428 | Heart failure | - |
| ICD-10 | I20 | Angina pectoris | 131296 |
| | I21 | Acute myocardial infarction | 131298 |
| | I22 | Subsequent myocardial infarction | 131300 |
| | I24 | Other acute ischaemic heart diseases | 131304 |
| | I25 | Chronic ischaemic heart disease | 131306 |
| | I42 | Cardiomyopathy | 131338 |
| | I44 | Atrioventricular and left bundle-branch block | 131342 |
| | I46 | Cardiac arrest | 131346 |
| | I47 | Paroxysmal tachycardia | 131348 |
| | I48 | Atrial fibrillation and flutter | 131350 |
| | I49 | Other cardiac arrhythmias | 131352 |
| | I50 | Heart failure | 131354 |

Table 2. Biomarker differences for participants with and without incident cardiovascular disease. IQR: inter-quartile range, LV: left ventricular, EF: ejection fraction, ED, end diastolic; ES, end systolic. All variables are distributed non-normally and compared using the Mann-Whitney U-test.

| | No incident CVD | | Incident CVD | | |
|---|---|---|---|---|---|
| **Outcome** | **Median (IQR)** | **N (%)** | **Median (IQR)** | **N (%)** | **p-value** |
| **Cohort: no type 2 diabetes** | | | | | |
| Ventricular rate, bpm | 61 (55-68) | 1683 (100) | 62 (55.2-72.2) | 98 (100) | 0.218 |
| QRS duration, ms | 88 (82-96) | 1683 (100) | 90 (82-98) | 98 (100) | 0.078 |
| QTc interval, ms | 420 (404-436) | 1683 (100) | 422 (413-444) | 98 (100) | 0.100 |
| T-wave offset, ms | 854 (834-874) | 1683 (100) | 848 (824-872) | 98 (100) | 0.140 |
| J-point amplitude (V3), mV | -0.015 (-0.044-0.019) | 1683 (100) | 0 (-0.0387-0.034) | 98 (100) | 0.031 |
| T-wave amplitude (V3), mV | 0.366 (0.229-0.533) | 1668 (99.1) | 0.407 (0.257-0.59) | 98 (100) | 0.076 |
| J-point amplitude (aVL), mV | 0.009 (-0.01-0.029) | 1683 (100) | 0.009 (-0.015-0.029) | 98 (100) | 0.570 |
| T-wave amplitude (aVL), mV | 0.112 (0.058-0.166) | 1623 (96.4) | 0.092 (0.019-0.156) | 95 (96.9) | 0.049 |
| Sokolow-Lyon index, mm | 20.1 (16.2-24.6) | 1564 (92.9) | 23.4 (17.7-27.1) | 86 (87.8) | 0.005 |
| LVEF, % | 56 (52-59) | 1445 (85.9) | 56 (50-60.2) | 88 (89.8) | 0.682 |
| LV ED volume, ml | 140 (119-162) | 1445 (85.9) | 142 (120-167) | 88 (89.8) | 0.437 |
| LV ES volume, ml | 61 (51-73) | 1445 (85.9) | 61 (50-78.2) | 88 (89.8) | 0.447 |
| LV stroke volume, ml | 78 (65-90) | 1445 (85.9) | 79 (65.8-88.2) | 88 (89.8) | 0.953 |
| Cardiac output, ml | 4.7 (4-5.6) | 1445 (85.9) | 4.85 (4.17-5.4) | 88 (89.8) | 0.483 |
| LV mass, g | 91.9 (76.1-108) | 1425 (84.7) | 101 (82.6-118) | 93 (94.9) | 0.001 |
| LV global average wall thickness, mm | 5.92 (5.39-6.44) | 1423 (84.6) | 6.29 (5.55-6.87) | 93 (94.9) | 0.000 |
| **Cohort: type 2 diabetes** | | | | | |
| Ventricular rate, bpm | 66 (59-74) | 1683 (100) | 66 (57-75.8) | 98 (100) | 0.913 |
| QRS duration, ms | 86 (80-94) | 1683 (100) | 88 (82-98) | 98 (100) | 0.032 |
| QTc interval, ms | 424 (408-440) | 1683 (100) | 424 (402-441) | 98 (100) | 0.682 |
| T-wave offset, ms | 842 (820-864) | 1683 (100) | 839 (816-862) | 98 (100) | 0.325 |
| J-point amplitude (V3), mV | -0.015 (-0.044-0.019) | 1683 (100) | -0.005 (-0.044-0.028) | 98 (100) | 0.472 |
| T-wave amplitude (V3), mV | 0.332 (0.205-0.488) | 1667 (99) | 0.395 (0.191-0.501) | 98 (100) | 0.343 |
| J-point amplitude (aVL), mV | 0.004 (-0.015-0.029) | 1683 (100) | 0.0065 (-0.01-0.034) | 98 (100) | 0.395 |
| T-wave amplitude (aVL), mV | 0.097 (0.043-0.146) | 1582 (94) | 0.117 (0.022-0.185) | 87 (88.8) | 0.294 |
| Sokolow-Lyon index, mm | 19.1 (15.2-23.4) | 1526 (90.7) | 20.5 (16.2-25.3) | 86 (87.8) | 0.094 |
| LVEF, % | 56 (51-59) | 1412 (83.9) | 55 (50-59) | 89 (90.8) | 0.118 |
| LV ED volume, ml | 130 (109-155) | 1412 (83.9) | 136 (116-155) | 89 (90.8) | 0.296 |
| LV ES volume, ml | 58 (47-71) | 1412 (83.9) | 62 (51-74) | 89 (90.8) | 0.106 |
| LV stroke volume, ml | 72 (60-85) | 1412 (83.9) | 72 (58-83) | 89 (90.8) | 0.808 |
| Cardiac output, ml | 4.7 (4-5.5) | 1412 (83.9) | 4.8 (4-5.4) | 89 (90.8) | 0.665 |
| LV mass, g | 90.9 (76.2-108) | 1426 (84.7) | 99.6 (85.8-107) | 92 (93.9) | 0.006 |
| LV global average wall thickness, mm | 6.08 (5.56-6.6) | 1424 (84.6) | 6.28 (5.86-6.81) | 92 (93.9) | 0.006 |

Table 3. Logistic regression models used to quantify the association of selected biomarker predictors with incident cardiovascular disease (single binary outcome). Models are adjusted sequentially for different types of confounding factors. Socio-demographic factors include age, sex, ethnicity; lifestyle factors include body mass index (BMI), smoking; clinical factors include diastolic blood pressure, total cholesterol, triglycerides, C-reactive protein, anti-hypertensive medication and insulin.

| | **No type 2 diabetes** | | | | | | | | | | | |
|---|---|---|---|---|---|---|---|---|---|---|---|---|
| | **Model 0** Unadjusted | | | **Model 1** Adjusted for socio-demographic factors | | | **Model 2** Additionally adjusted for lifestyle factors | | | **Model 3** Additionally adjusted for clinical factors | | |
| **Predictor** | **N** | **Coefficient (95% CI)** | **p-value** | **N** | **Coefficient (95% CI)** | **p-value** | **N** | **Coefficient (95% CI)** | **p-value** | **N** | **Coefficient (95% CI)** | **p-value** |
| QRS duration, ms | 1781 | 0.016 (0.004-0.027) | 0.007 | 1781 | 0.015 (0.002-0.027) | 0.019 | 1762 | 0.014 (0.002-0.026) | 0.022 | 1464 | 0.0137 (0-0.027) | 0.046 |
| Sokolow-Lyon index, mm | 1650 | 0.049 (0.019-0.079) | 0.001 | 1650 | 0.049 (0.017-0.081) | 0.003 | 1633 | 0.051 (0.017-0.084) | 0.003 | 1364 | 0.051 (0.015-0.086) | 0.005 |
| LV mass, g | 1518 | 0.017 (0.008-0.025) | <0.001 | 1518 | 0.0248 (0.014-0.036) | <0.001 | 1501 | 0.027 (0.015-0.039) | <0.001 | 1297 | 0.029 (0.017-0.042) | <0.001 |
| LV global average wall thickness, mm | 1516 | 0.508 (0.253-0.76) | <0.001 | 1516 | 0.623 (0.316-0.926) | <0.001 | 1499 | 0.707 (0.357-1.05) | <0.001 | 1295 | 0.677 (0.293-1.06) | <0.001 |

| | **Type 2 diabetes** | | | | | | | | | | | |
|---|---|---|---|---|---|---|---|---|---|---|---|---|
| | **Model 0** Unadjusted | | | **Model 1** Adjusted for socio-demographic factors | | | **Model 2** Additionally adjusted for lifestyle factors | | | **Model 3** Additionally adjusted for clinical factors | | |
| **Predictor** | **N** | **Coefficient (95% CI)** | **p-value** | **N** | **Coefficient (95% CI)** | **p-value** | **N** | **Coefficient (95% CI)** | **p-value** | **N** | **Coefficient (95% CI)** | **p-value** |
| QRS duration, ms | 1781 | 0.0143 (0.002-0.026) | 0.020 | 1781 | 0.013 (0-0.025) | 0.045 | 1761 | 0.013 (0-0.025) | 0.042 | 1456 | 0.009 (-0.007-0.023) | 0.263 |
| Sokolow-Lyon index, mm | 1612 | 0.031 (-0.001-0.062) | 0.053 | 1612 | 0.030 (-0.003-0.062) | 0.071 | 1592 | 0.0363 (0.002-0.069) | 0.032 | 1335 | 0.0209 (-0.018-0.058) | 0.280 |
| LV mass, g | 1518 | 0.0103 (0.001-0.019) | 0.022 | 1518 | 0.0136 (0.003-0.024) | 0.013 | 1500 | 0.016 (0.003-0.027) | 0.011 | 1285 | 0.019 (0.006-0.032) | 0.003 |
| LV global average wall thickness, mm | 1516 | 0.354 (0.091-0.614) | 0.008 | 1516 | 0.413 (0.107-0.71) | 0.007 | 1498 | 0.457 (0.115-0.79) | 0.008 | 1283 | 0.561 (0.191-0.923) | 0.003 |

Table 4. Subgroup analysis: changes in biomarkers within the glycaemic spectrum. Multivariate multiple linear regression models to quantify the association of HbA1c with selected ECG and CMR-derived biomarkers. Models are adjusted sequentially for different types of confounding factors. Socio-demographic factors include age, sex, ethnicity; lifestyle factors include body mass index (BMI), smoking; clinical factors include diastolic blood pressure, total cholesterol, triglycerides, C-reactive protein, anti-hypertensive medication and insulin.

| Outcome | **Model 0** Unadjusted **N = 1170** | | **Model 1** Adjusted for socio-demographic factors **N = 1170** | | **Model 2** Additionally adjusted for lifestyle factors **N = 1154** | | **Model 3** Additionally adjusted for clinical factors **N = 1014** | |
|---|---|---|---|---|---|---|---|---|
| | **Coefficient (95% CI)** | **p-value** | **Coefficient (95% CI)** | **p-value** | **Coefficient (95% CI)** | **p-value** | **Coefficient (95% CI)** | **p-value** |
| **ECG** | | | | | | | | |
| Ventricular rate, bpm | 0.095 (0.037-0.153) | 0.001 | 0.094 (0.036-0.151) | 0.001 | 0.097 (0.040-0.154) | 0.001 | 0.12 (0.054-0.187) | 0.000 |
| QRS duration, ms | -0.020 (-0.088-0.047) | 0.557 | -0.009 (-0.073-0.056) | 0.79 | -0.0114 (-0.076-0.054) | 0.731 | 0.011 (-0.063-0.086) | 0.768 |
| QTc interval, ms | 0.012 (-0.102-0.126) | 0.835 | 0.004 (-0.105-0.114) | 0.937 | 0.009 (-0.100-0.118) | 0.871 | 0.026 (-0.106-0.157) | 0.702 |
| T-wave offset, ms | -0.26 (-0.425-(-0.096)) | 0.002 | -0.267 (-0.429-(-0.105)) | 0.001 | -0.279 (-0.441-(-0.116)) | 0.001 | -0.33 (-0.522-(-0.138)) | 0.001 |
| T-wave amplitude (V3), mV | 0.0001 (-0.001-0.001) | 0.859 | 0.0003 (-0.001-0.001) | 0.59 | 0.0002 (-0.001-0.001) | 0.642 | -0.001 (-0.002-0.000) | 0.19 |
| T-wave amplitude (aVL), mV | -0.001 (-0.001-0.000) | 0.012 | -0.001 (-0.001-0.000) | 0.013 | -0.001 (-0.001-0.000) | 0.017 | 0.000 (-0.001-0.000) | 0.151 |
| J-point amplitude (V3), mV | 0.000 (-0.0001-0.0003) | 0.547 | 0.0001 (-0.0001-0.0004) | 0.384 | 0.000 (-0.0001-0.0004) | 0.466 | 0.000 (-0.0003-0.0004) | 0.816 |
| J-point amplitude (aVL), mV | 0.000 (-0.0002-0.0002) | 0.869 | 0.000 (-0.0002-0.0002) | 0.817 | 0.000 (-0.0002-0.0002) | 0.925 | 0.000 (-0.0002-0.0002) | 0.784 |
| Sokolow-Lyon index, mm | 0.010 (-0.023-0.042) | 0.555 | 0.010 (-0.022-0.042) | 0.558 | 0.008 (-0.024-0.039) | 0.638 | 0.006 (-0.031-0.044) | 0.736 |
| **CMR** | | | | | | | | |
| LV ED volume, ml | -0.363 (-0.837-0.111) | 0.133 | -0.347 (-0.814-0.121) | 0.146 | -0.323 (-0.797-0.151) | 0.182 | -0.256 (-0.852-0.34) | 0.4 |
| LV stroke volume, ml | -0.135 (-0.256-(-0.014)) | 0.029 | -0.132 (-0.247-(-0.018)) | 0.023 | -0.122 (-0.237-(-0.007)) | 0.038 | -0.104 (-0.243-0.036) | 0.145 |
| LV global average wall thickness, mm | -0.002 (-0.006-0.002) | 0.378 | -0.001 (-0.005-0.002) | 0.408 | -0.0003 (-0.003-0.003) | 0.86 | 0.001 (-0.003-0.004) | 0.607 |

Table 5. Subgroup analysis: females versus males. IQR: inter-quartile range, ECG: electrocardiogram, CMR: cardiac magnetic resonance, LV: left ventricular, EF: ejection fraction, ED: end-diastolic, ES: end-systolic. All continuous variables are distributed non-normally and compared using the Mann-Whitney U-test.

| | **No type 2 diabetes** | | **Type 2 diabetes** | | |
|---|---|---|---|---|---|
| | **Median (IQR)** | **N (%)** | **Median (IQR)** | **N (%)** | **p-value** |
| **Cohort: females** | | | | | |
| Ventricular rate, bpm | 62 (57-70) | 648 (100) | 67 (60-74) | 648 (100) | <0.001 |
| QRS duration, ms | 82 (76-90) | 648 (100) | 82 (76-88) | 648 (100) | 0.048 |
| QTc interval, ms | 429 (413-443) | 648 (100) | 433 (417-447) | 648 (100) | 0.001 |
| T-wave offset, ms | 858 (838-876) | 648 (100) | 850 (826-870) | 648 (100) | <0.001 |
| J-point amplitude (V3), mV | -0.03 (-0.054-0) | 648 (100) | -0.025 (-0.054-0.004) | 648 (100) | 0.208 |
| T-wave amplitude (V3), mV | 0.263 (0.156-0.375) | 638 (98.5) | 0.239 (0.141-0.341) | 635 (98) | 0.040 |
| J-point amplitude (aVL), mV | 0.004 (-0.015-0.029) | 648 (100) | 0.004 (-0.015-0.029) | 648 (100) | 0.381 |
| T-wave amplitude (aVL), mV | 0.097 (0.043-0.156) | 615 (94.9) | 0.087 (0.007-0.132) | 600 (92.6) | 0.006 |
| Sokolow-Lyon index, mm | 18.1 (14.8-22.1) | 609 (94) | 17.4 (14.2-21.5) | 584 (90.1) | 0.019 |
| LVEF, % | 57 (54-61) | 564 (87) | 57 (53-61) | 551 (85) | 0.526 |
| LV ED volume, ml | 120 (104-138) | 564 (87) | 113 (97-130) | 551 (85) | <0.001 |
| LV ES volume, ml | 51 (43-60) | 564 (87) | 48 (41-57.5) | 551 (85) | <0.001 |
| LV stroke volume, ml | 68 (58-79) | 564 (87) | 64 (54-74) | 551 (85) | <0.001 |
| Cardiac output, ml | 4.2 (3.7-4.8) | 564 (87) | 4.3 (3.6-4.9) | 551 (85) | 0.852 |
| LV mass, g | 72.8 (63.9-81.2) | 562 (86.7) | 73.2 (65.4-84.2) | 556 (85.8) | 0.062 |
| LV global average wall thickness, mm | 5.31 (4.98-5.67) | 561 (86.6) | 5.55 (5.17-5.94) | 556 (85.8) | <0.001 |
| **Cohort: males** | | | | | |
| Ventricular rate, bpm | 60 (54-67) | 1133 (100) | 65 (58-73) | 1133 (100) | <0.001 |
| QRS duration, ms | 90 (84-98) | 1133 (100) | 90 (84-98) | 1133 (100) | 0.018 |
| QTc interval, ms | 416 (401-431) | 1133 (100) | 419 (404-434) | 1133 (100) | 0.004 |
| T-wave offset, ms | 850 (832-872) | 1133 (100) | 838 (814-860) | 1133 (100) | <0.001 |
| J-point amplitude (V3), mV | -0.005 (-0.035-0.034) | 1133 (100) | -0.005 (-0.035-0.034) | 1133 (100) | 0.532 |
| T-wave amplitude (V3), mV | 0.434 (0.302-0.61) | 1128 (99.6) | 0.395 (0.258-0.55) | 1130 (99.7) | <0.001 |
| J-point amplitude (aVL), mV | 0.009 (-0.01-0.034) | 1133 (100) | 0.009 (-0.015-0.029) | 1133 (100) | 0.017 |
| T-wave amplitude (aVL), mV | 0.117 (0.063-0.17) | 1103 (97.4) | 0.102 (0.048-0.156) | 1069 (94.4) | <0.001 |
| Sokolow-Lyon index, mm | 21.4 (17.2-25.9) | 1041 (91.9) | 20.4 (16-24.5) | 1028 (90.7) | <0.001 |
| LVEF, % | 55 (51-59) | 969 (85.5) | 54 (50-58) | 950 (83.8) | 0.072 |
| LV ED volume, ml | 152 (134-173) | 969 (85.5) | 142 (122-165) | 950 (83.8) | <0.001 |
| LV ES volume, ml | 68 (58-80) | 969 (85.5) | 65 (53-77) | 950 (83.8) | <0.001 |
| LV stroke volume, ml | 83 (72-96) | 969 (85.5) | 77 (64-90) | 950 (83.8) | <0.001 |
| Cardiac output, ml | 5 (4.4-5.8) | 969 (85.5) | 5 (4.2-5.8) | 950 (83.8) | <0.001 |
| LV mass, g | 103 (92.6-114) | 956 (84.4) | 103 (89.7-115) | 962 (84.9) | <0.001 |
| LV global average wall thickness, mm | 6.26 (5.86-6.73) | 955 (84.3) | 6.37 (5.97-6.84) | 960 (84.7) | <0.001 |

Table 6. Subgroup analysis: white versus non-white participants. IQR: inter-quartile range, ECG: electrocardiogram, CMR: cardiac magnetic resonance, LV: left ventricular, EF: ejection fraction, ED: end-diastolic, ES: end-systolic. All continuous variables are distributed non-normally and compared using the Mann-Whitney U-test.

| | **No type 2 diabetes** | | **Type 2 diabetes** | | |
|---|---|---|---|---|---|
| | **Median (IQR)** | **N (%)** | **Median (IQR)** | **N (%)** | **p-value** |
| **Cohort: non-white ethnic background** | | | | | |
| Ventricular rate, bpm | 59.5 (56.5-66) | 48 (100) | 67 (59-76) | 141 (100) | <0.001 |
| QRS duration, ms | 84 (80-90.5) | 48 (100) | 84 (78-92) | 141 (100) | 0.871 |
| QTc interval, ms | 415 (397-432) | 48 (100) | 420 (406-440) | 141 (100) | 0.141 |
| T-wave offset, ms | 851 (833-867) | 48 (100) | 836 (812-860) | 141 (100) | 0.004 |
| J-point amplitude (V3), mV | -0.008 (-0.045-0.043) | 48 (100) | 0 (-0.04-0.048) | 141 (100) | 0.406 |
| T-wave amplitude (V3), mV | 0.375 (0.229-0.556) | 47 (97.9) | 0.312 (0.194-0.463) | 140 (99.3) | 0.188 |
| J-point amplitude (aVL), mV | 0.024 (0.003-0.048) | 48 (100) | 0.009 (-0.015-0.034) | 141 (100) | 0.027 |
| T-wave amplitude (aVL), mV | 0.117 (0.085-0.188) | 47 (97.9) | 0.112 (0.063-0.17) | 135 (95.7) | 0.130 |
| Sokolow-Lyon index, mm | 20.9 (17.4-25.9) | 40 (83.3) | 20.4 (15.5-25.7) | 124 (87.9) | 0.368 |
| LVEF, % | 56 (51-60) | 38 (79.2) | 56 (53-60) | 112 (79.4) | 0.898 |
| LV ED volume, ml | 124 (115-146) | 38 (79.2) | 119 (97.8-139) | 112 (79.4) | 0.146 |
| LV ES volume, ml | 56.5 (47-71) | 38 (79.2) | 51 (43-66) | 112 (79.4) | 0.156 |
| LV stroke volume, ml | 68.5 (62-83.5) | 38 (79.2) | 66 (54-78) | 112 (79.4) | 0.143 |
| Cardiac output, ml | 4.45 (3.73-5.08) | 38 (79.2) | 4.5 (3.6-5.3) | 112 (79.4) | 0.782 |
| LV mass, g | 93.3 (81.2-103) | 38 (79.2) | 86.5 (70.3-103) | 110 (78) | 0.285 |
| LV global average wall thickness, mm | 6.26 (5.51-6.68) | 38 (79.2) | 6.01 (5.51-6.55) | 110 (78) | 0.484 |
| **Cohort: white ethnic background** | | | | | |
| Ventricular rate, bpm | 61 (55-68) | 1733 (100) | 66 (59-74) | 1640 (100) | <0.001 |
| QRS duration, ms | 88 (82-96) | 1733 (100) | 86 (80-96) | 1640 (100) | 0.013 |
| QTc interval, ms | 421 (405-436) | 1733 (100) | 424 (408-440) | 1640 (100) | <0.001 |
| T-wave offset, ms | 854 (834-874) | 1733 (100) | 842 (820-864) | 1640 (100) | <0.001 |
| J-point amplitude (V3), mV | -0.015 (-0.044-0.019) | 1733 (100) | -0.015 (-0.044-0.019) | 1640 (100) | 0.699 |
| T-wave amplitude (V3), mV | 0.366 (0.229-0.537) | 1719 (99.2) | 0.332 (0.205-0.488) | 1625 (99.1) | <0.001 |
| J-point amplitude (aVL), mV | 0.009 (-0.01-0.029) | 1733 (100) | 0.004 (-0.015-0.029) | 1640 (100) | 0.021 |
| T-wave amplitude (aVL), mV | 0.107 (0.053-0.166) | 1671 (96.4) | 0.097 (0.039-0.146) | 1534 (93.5) | <0.001 |
| Sokolow-Lyon index, mm | 20.1 (16.2-24.7) | 1610 (92.9) | 19.1 (15.2-23.4) | 1488 (90.7) | <0.001 |
| LVEF, % | 56 (52-59) | 1495 (86.3) | 55 (51-59) | 1389 (84.7) | 0.041 |
| LV ED volume, ml | 141 (119-163) | 1495 (86.3) | 131 (110-156) | 1389 (84.7) | <0.001 |
| LV ES volume, ml | 61 (51-74) | 1495 (86.3) | 58 (48-72) | 1389 (84.7) | <0.001 |
| LV stroke volume, ml | 78 (65-90) | 1495 (86.3) | 72 (60-85) | 1389 (84.7) | <0.001 |
| Cardiac output, ml | 4.7 (4.1-5.6) | 1495 (86.3) | 4.7 (4-5.5) | 1389 (84.7) | 0.730 |
| LV mass, g | 92.5 (76.3-109) | 1480 (85.4) | 91.6 (77.2-108) | 1408 (85.9) | 0.555 |
| LV global average wall thickness, mm | 5.92 (5.4-6.46) | 1478 (85.3) | 6.09 (5.6-6.62) | 1406 (85.7) | <0.001 |
